# Efficacy of a bivalent (D614 + B.1.351) SARS-CoV-2 Protein Vaccine

**DOI:** 10.1101/2022.12.05.22282933

**Authors:** Gustavo H. Dayan, Nadine Rouphael, Stephen R. Walsh, Aiying Chen, Nicole Grunenberg, Mary Allen, Johannes Antony, Kwaku Poku Asante, Amit Suresh Bhate, Tatiana Beresnev, Matthew I Bonaparte, Maria Angeles Ceregido, Dmytro Dobrianskyi, Bo Fu, Marie-Helene Grillet, Maryam Keshtkar-Jahromi, Michal Juraska, Jia Jin Kee, Hannah Kibuuka, Marguerite Koutsoukos, Roger Masotti, Nelson L. Michael, Humberto Reynales, Merlin L. Robb, Sandra M. Villagómez Martínez, Fredrick Sawe, Lode Schuerman, Tina Tong, John Treanor, T. Anh Wartel, Carlos A. Diazgranados, Roman M. Chicz, Sanjay Gurunathan, Stephen Savarino, Saranya Sridhar, the VAT00008 study team

**Author notes:** **Corresponding Author:** Gustavo H. Dayan. Authors contributed equally to the study. Affiliation at time of study.

## Abstract

**Background:** COVID-19 vaccines with alternative strain compositions are needed to provide broad protection against newly emergent SARS-CoV-2 variants of concern.

**Methods:** We conducted a global Phase 3, multi-stage efficacy study (NCT04904549) among adults aged ≥18 years. Participants were randomized 1:1 to receive two intramuscular injections 21 days apart of a bivalent SARS-CoV-2 recombinant protein vaccine with AS03-adjuvant (5 μg of ancestral (D614) and 5 μg of B.1.351 [beta] variant spike protein) or placebo. Symptomatic COVID-19 was defined as laboratory-confirmed COVID-19 with COVID-19-like illness (CLI) symptoms. The primary efficacy endpoint was the prevention of symptomatic COVID-19 ≥14 days after the second injection (post-dose 2 [PD2]).

**Results:** Between 19 Oct 2021 and 15 Feb 2022, 12,924 participants received ≥1 study injection. 75% of participants were SARS-CoV-2 non-naïve. 11,416 participants received both study injections (efficacy-evaluable population [vaccine, n=5,736; placebo, n=5,680]). Up to 15 March 2022, 121 symptomatic COVID-19 cases were reported (32 in the vaccine group and 89 in the placebo group) ≥14 days PD2 with a vaccine efficacy (VE) of 64.7% (95% confidence interval [CI] 46.6; 77.2%). VE was 75.1% (95% CI 56.3; 86.6%) in non-naïve and 30.9% (95% CI -39.3; 66.7%) in naïve participants. Viral genome sequencing identified the infecting strain in 68 cases (Omicron [BA.1 and BA.2 subvariants]: 63; Delta: 4; Omicron and Delta: 1). The vaccine was well-tolerated and had an acceptable safety profile.

**Conclusions:** A bivalent vaccine conferred heterologous protection against symptomatic infection with newly emergent Omicron (BA.1 and BA.2) in non-naïve adults 18–59 years of age.

**ClinicalTrials.gov:** NCT04904549

## Introduction

First-generation COVID-19 vaccines were developed using the Spike (S) sequence from the SARS-CoV-2 ancestral Wuhan-Hu-1 (D614) strain.^1^ However, these vaccines are less effective against new emergent SARS-CoV-2 variants of concern (VOCs; including Omicron [BA.1, BA.2, BA.4 and BA.5] variants) ^2-7^ Vaccines with variant strains have been developed to provide cross-protection against emerging variants. One strategy for variant vaccine composition is inclusion of the prevalent circulating strain, with mRNA Omicron-containing bivalent vaccines authorized as boosters based on demonstrated induction of antibodies to circulating Omicron variants.^8,9^ However, there are no data on whether an alternative non-Omicron variant vaccine provides cross-protective efficacy against Omicron variants.

Sanofi and GSK have developed a bivalent vaccine containing stabilized SARS-CoV-2 pre-fusion S proteins from both the ancestral D614 and the Beta (B.1.351) variant, with the GSK AS03 adjuvant system (CoV2 preS dTM-AS03 [D614 + B.1.351]). This bivalent vaccine is being evaluated as a two-injection primary series in previously unvaccinated individuals and as a booster vaccine based on preclinical studies showing induction of cross-neutralizing antibody responses against a broad panel of VOCs.^10,11^ For the first time, we describe the clinical efficacy and safety of two injections of the bivalent vaccine as a primary series during a period of Omicron circulation.

## Methods

### Trial Design

This Phase 3, parallel, international, randomized, modified double-blind, placebo-controlled study was designed as a multi-stage platform trial with two stages (NCT04904549). Stage 1 evaluated the efficacy of the prototype vaccine, containing the ancestral D614 recombinant S protein (CoV2 preS dTM-AS03 [D614]) (manuscript in preparation). Stage 2, reported here, evaluated the efficacy and safety of a primary series of two injections of the bivalent vaccine, administered 21 days apart. Stage 2 was conducted in 54 clinical research centers across eight countries: Colombia, Ghana, India, Kenya, Mexico, Nepal, Uganda and Ukraine **(Supplementary Appendix Section 1.1)**. Participant enrollment started on 19 October 2021 and finished on 15 February 2022. Eligible participants were randomized 1:1 to receive either the bivalent vaccine or placebo (saline) (**Supplementary Appendix 1.2**).

The study was conducted in compliance with the International Conference on Harmonization (ICH) guidelines for Good Clinical Practice and the principles of the Declaration of Helsinki. The protocol and amendments were approved by applicable Independent Ethics Committees/Institutional Review Boards and per local regulations. Approval was received by the following Independent Ethics Committees/Institutional Review Boards. Colombia: Comité de Ética en Investigación CAIMED (approved); Comité de Ética en Investigación de la Fundación del Caribe para la Investigación Biomédica (approved); Comité de Ética en Investigación VITA (approved); Corporación Científica Pediátrica Comité de Etica en Investigación Biomédica (approved); Comité de Ética en investigación de la División Ciencias de la Salud de la Universidad del Norte (approved); Comité de Ética en Investigación Clínica de la Costa (approved). Ghana: Kwame Nkrumah University of Science and Technology Committee on Human Research, Publication and Ethics (approved); Kintampo Health Research Centre Institutional Ethics Committee (approved); Navrongo Health Research Centre Institutional Review Board (approved); Ghana Health Service Ethics Review Committee (approved). India: Institute of Medical Sciences and Sum Hospital Institutional Ethics Committee (approved); Jawahar Lal Nehru Medical College Institutional Ethics Committee (approved); Vidharba Institute of Medical Sciences - Nagpur Institutional Ethics Committee (approved); Jeevan Rekha Hospital Institutional Ethics Committee (approved); SRM Medical College Hospital and Research Centre Institutional Ethics Committee (approved); Prakhar Hospital Institutional Ethics Committee (approved); All India Institute of Medical Sciences Patna Institutional Ethics Committee (approved); Aartham Ethics Committee - Aartham Multi Super Speciality Hospital (approved); Maharaja Agrasen Superspeciality Hospital Institutional Ethics Committee (approved). Kenya: Kenya Medical Research Scientific & Institute Ethics Review Unit (approved); Kenyatta National Hospital - University of Nairobi - College of Health Sciences Ethics Review Committee (approved); The Aga Khan University - Nairobi Institutional Ethics Review Committee (approved); Institutional Review Ethics Committee MOI University College of Health - MOI Teaching and Referral Hospital (approved); National Commission for Science Technology and Innovation (approved). Mexico: Comité de Ética en Investigación de la Dirección de Investigación del Instituto Nacional de Pediatría; Comité de Ética en Investigación de Médica Sur (approved); Comité de Ética en Investigación Hospital Aranda de la Parra (approved); Comité de Ética en Investigación Hospital Civil de Guadalajara - Sub-Dirección de Enseñanza e Investigación (approved); Comité de Ética en Investigación de Investigación Biomédica para el Desarrollo de Fármacos (approved). Nepal: Tribhuvan University Institute of Medicine Institutional Review Committee (approved); Kathmandu University School of Medical Sciences Institutional Review Board (approved); Nepalgunj Medical College Teaching Hospital Institutional Review Committee (approved). Uganda: Uganda Virus Research Institute (approved); London School of Hygiene and Tropical Medicine Research Ethics Committee (approved). Ukraine: Blagomed Medical Clinic LLC Ethics Commission (approved); Edelweiss Medics LLC (approved); Center of Family Medicine Plus LLC Ethical Committee (approved); Medbud Clinic LLC Ethical Committee (approved). All participants provided informed consent. An independent data and safety monitoring board^12^ provided study oversight and reviewed unblinded data.

## Participants

Adults aged ≥18 years who had not received a prior COVID-19 vaccine were eligible for inclusion; full details of the inclusion and exclusion criteria are reported in the **Supplementary Appendix Section 1.3**. Efforts were made to make participants aware of the availability of approved/authorized COVID-19 vaccines (**Supplementary Appendix Section 1.4**). Participants with a potentially high risk for severe COVID-19 (**Supplementary Appendix Section 1.5**) and other subpopulations at risk of COVID-19 infection, including ethnic and racial minorities, were included.

## Interventions and assessments

The recombinant protein antigen CoV2 preS dTM and the AS03 Adjuvant System (GSK Vaccines, Rixensart, Belgium) have been described previously.^13-15^ Briefly, CoV2 preS dTM, stabilized in its prefusion form, is produced using the baculovirus expression system technology. Each 0.5 mL injection of the bivalent vaccine formulation contained 5 μg of the ancestral D614 and 5 μg of the B.1.351 variant Spike protein antigen. The injection protocol is reported in **Supplementary Appendix Section 1.6**. Vaccinations were administered on study days 1 and 22 by intramuscular injection into the deltoid region by qualified and trained personnel.

Blood samples and nasopharyngeal swabs were collected before each vaccination to establish whether participants had previous or ongoing SARS-CoV-2 infection (naïve or non-naïve). Testing procedures and criteria for determination of prior SARS-CoV-2 infection are described in the **Supplementary Appendix Section 1.7**.

Surveillance for COVID-19-like illness (CLI) was both active and passive: participants were contacted once a week to determine whether they had any symptoms of a CLI (**Supplementary Appendix Section 1.8**) or if they had a positive COVID-19 test from another source at any time during the study. In the event of CLI symptoms, nasopharyngeal and anterior nasal swabs were collected at the participant’s first visit after symptom onset and 2–4 days later for virological confirmation using NAAT (**Supplementary Appendix Section 1.9**). An independent adjudication committee reviewed potential cases to determine whether the case definitions for symptomatic and/or severe COVID-19 were met. Viral genomic sequencing was performed on respiratory samples from the cases to identify the SARS-CoV-2 variant, as previously described.^16, 17^

### Efficacy endpoints

The primary efficacy objective was to assess in all participants, regardless of prior infection, the clinical efficacy of the bivalent vaccine for prevention of symptomatic COVID-19 ≥14 days after the second injection (post-dose 2 [PD2]). Secondary efficacy endpoints included the occurrence of symptomatic disease in naïve and non-naïve individuals; and severe, moderate or worse, or hospitalized COVID-19 ≥14 days PD2 in all participants and according to prior infection status. Additional reported analyses and all endpoints are defined in **Supplementary Appendix Sections 1.10** and **1.11**.

### Safety

Participants were directed to report any adverse events (AEs) during their study visits or during any follow-up contact with the investigators. Safety data were collected from all participants receiving at least one injection of the study vaccine or placebo (**Supplementary Appendix Section 1.12**) throughout the duration of the study. Solicited injection site reactions (SISRs) and solicited systemic reactions (SSRs) occurring within 7 days after each vaccination and non-serious unsolicited AEs occurring within 21 days after each vaccination were collected in a subset of approximately 4,000 participants (the first 4000 participants recruited [2000 in each arm], as well as all participants ≥60 years of age).

### Statistical Analyses

The data cut-off date for the analyses reported here was 15 March 2022. Calculations for determining this sample size are reported in **Supplementary Appendix Section 1.13**; descriptions of the analysis sets are reported in **Supplementary Section 1.14**.

Efficacy analyses were conducted on the modified full analysis set PD2 (mFAS-PD2), comprising participants who received both injections (excluding participants with onset of symptomatic COVID-19 between the first injection [post-dose 1 (PD1)] and 14 days PD2) who did not meet any vaccine contraindications and did not discontinue the study within 14 days PD2. These participants were further divided based on prior infection status PD1 and PD2.

For the primary endpoint, the point estimate of vaccine efficacy (VE) was calculated based on the incidence rate per 1000 person-years per group in the mFAS-PD2 population, regardless of prior infection status. The primary objective was met if the VE point estimate was >50% and the lower bound of the confidence interval (CI) was >30%. Survival analyses (Kaplan-Meier curves with 95% CI) were also performed. Sensitivity analyses were conducted assuming that unsequenced cases were due to the Omicron variant, which was the prevalent variant circulating at the time of the study. Safety outcomes were assessed in the safety analysis set (SafAS), comprising all randomized participants who received ≥1 injection of study vaccine or placebo. Statistical analyses were performed using SAS® Version 9.4 or later.

## Results Participants

Between 19 October 2021 and 15 February 2022, 13,506 participants were randomized. Owing to the ongoing war in Ukraine, data completeness could not be confirmed for the four Ukrainian sites; therefore, none of the 504 participants from these sites were included in the main analyses, although sensitivity analyses including these data were performed.

In the current analysis, 13,002 participants were randomized to receive the study vaccine (n=6,512) or placebo (n=6,490) up to the cut-off date of 15 March 2022 (**Figure 1**). Of those, 414 participants (3.2%) discontinued the study, 89 of whom discontinued PD2 (**Supplementary Appendix Section 2.1**). The main analysis sets are presented in **Supplementary Appendix 2.2**.

**Figure 1:**
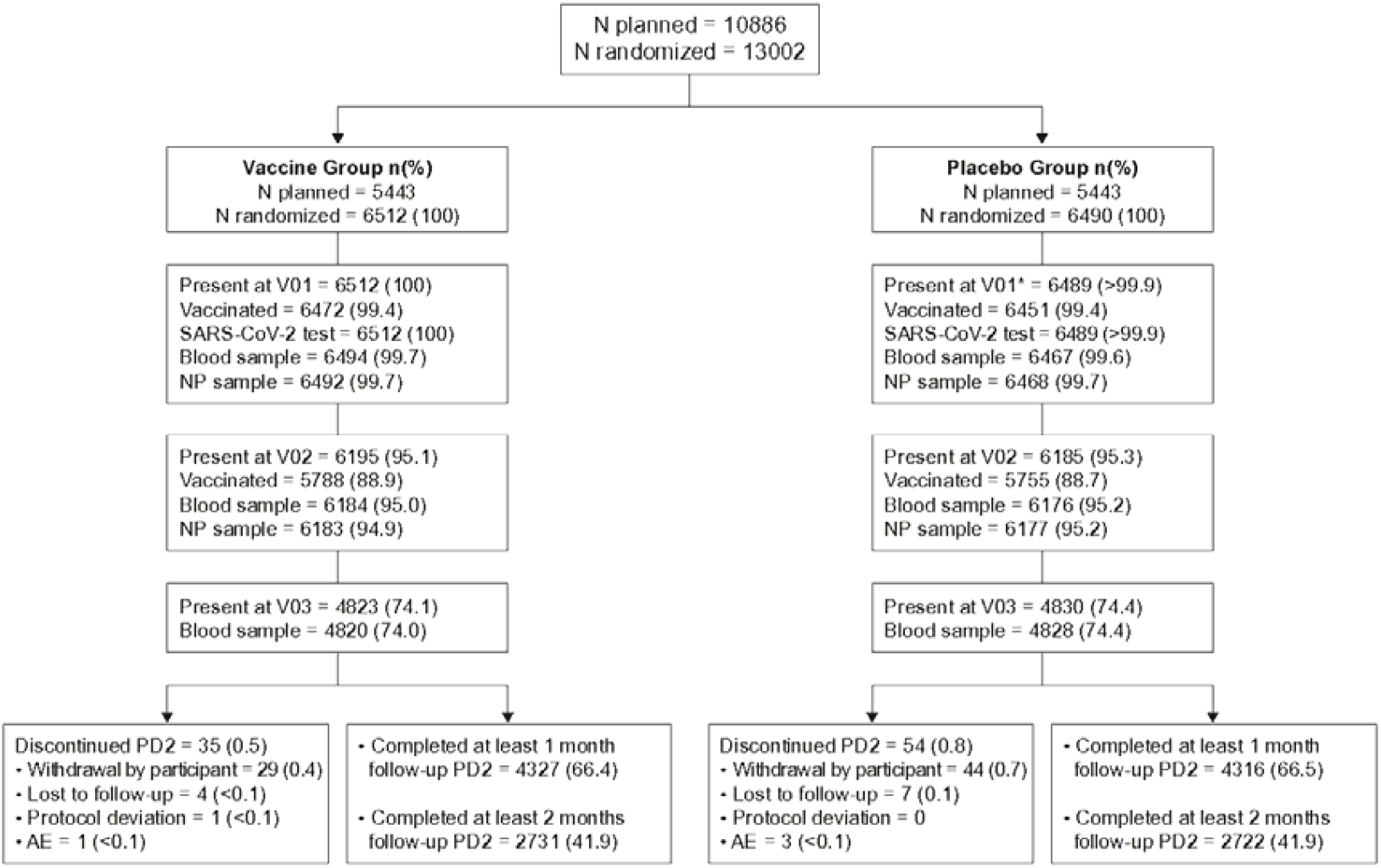
CONSORT diagram for patient flow through the study Data are presented as number (%). *V1 for one participant did not appear in the database during the data extraction dated 09 June 2022 because the site was entering additional data for V01 at the time the data extraction was performed. However, this participant was included in mFAS-PD1, mFAS-PD2, mFAS-PD2 Non-naïve-D01/D22 analysis sets because both V01 and V02 were performed. Abbreviations: AE, adverse event. NP, nasopharyngeal. PD2, post dose 2. V, visit.

A total of 12,924 participants received ≥1 study injection (SafAS), for whom demographic characteristics are reported based on first visit samples (**Table 1**). Patient demographics were comparable across treatment groups. The mean (SD) age was 36.1 (12.9) years and 58.4% were male (**Table 1**). 75% of participants had evidence of prior infection (non-naïve) at enrollment. High-risk medical conditions were present in 32.2% of participants (**Table 1** and **Supplementary Appendix Section 2.3**).

**Table 1:** Demographics and clinical characteristics at baseline in the participants who received at least one injection (SafAS)

|  | **Vaccine group**<br>**(N=6,472)** | **Placebo group**<br>**(N=6,450)** | **Total**<br>**(N=12,924\*)** |
| --- | --- | --- | --- |
| Sex, n (%) |  |  |  |
| Male | 3789 (58.5) | 3751 (58.2) | 7542 (58.4) |
| Female | 2683 (41.5) | 2699 (41.8) | 5382 (41.6) |
| Age, years |  |  |  |
| Mean (SD) | 36.1 (13.0) | 36.0 (12.9) | 36.1 (12.9) |
| Median (min; max) | 34.0 (18.0; 93.0) | 34.0 (18.0; 93.0) | 34.0 (18.0; 93.0) |
| Age categories, n (%) |  |  |  |
| 18-59 years | 6078 (93.9) | 6067 (94.1) | 12,147 (94.0) |
| ≥60 years | 394 (6.1) | 383 (5.9) | 777 (6.0) |
| BMI, mean (SD); median (Q1; Q3) | 23.8 (4.61);<br>22.9 (20.7; 25.8) | 23.8 (4.41)<br>22.9 (20.8; 25.8) | 23.8 (4.51)<br>22.9 (20.7; 25.8) |
| Race, n (%) |  |  |  |
| American Indian or Alaskan native | 408 (6.3) | 402 (6.2) | 811† (6.3) |
| Asian | 2562 (39.6) | 2567 (39.8) | 5129 (39.7) |
| Black or African American | 2873 (44.4) | 2854 (44.2) | 5727 (44.3) |
| White | 36 (0.6) | 38 (0.6) | 74 (0.6) |
| Multiracial | 5 (<0.1) | 6 (<0.1) | 11 (<0.1) |
| Not reported | 95 (1.5) | 82 (1.3) | 177 (1.4) |
| Ethnicity, n (%) |  |  |  |
| Hispanic or Latino | 1056 (16.3) | 1051 (16.3) | 2109† (16.3) |
| Not Hispanic or Latino | 5381 (83.1) | 5372 (83.3) | 10,753 (83.2) |
| Not reported | 15 (0.2) | 13 (0.2) | 28 (0.2) |
| Country, n (%) (%) |  |  |  |
| Mexico | 495 (7.6) | 493 (7.6) | 989 (7.7) |
| Colombia | 537 (8.3) | 532 (8.2) | 1070 (8.3) |
| India | 1661 (25.7) | 1672 (25.9) | 3333 (25.8) |
| Uganda | 212 (3.3) | 206 (3.2) | 418 (3.2) |
| Ghana | 597 (9.2) | 598 (9.3) | 1195 (9.2) |
| Kenya | 2066 (31.9) | 2052 (31.8) | 4118 (31.9) |
| Nepal | 904 (14.0) | 897 (13.9) | 1801 (13.9) |
| Prior SARS-CoV-2 infection, n (%) |  |  |  |
| Naïve at Day 1 | 588 (9.1) | 588 (9.1) | 1176 (9.1) |
| Non-naïve at Day 1 | 4860 (75.1) | 4831 (74.9) | 9693 (75.0) |
| Undetermined at Day 1 | 1024 (15.8) | 1031 (16.0) | 2055 (15.9) |
| Naïve at Day 22 | 333 (5.1) | 350 (5.4) | 683 (4.3) |
| Non-naïve at Day 22 | 5478 (84.6) | 5486 (85.1) | 10,966 (94.8) |
| Undetermined at Day 22 | 661 (10.2) | 614 (9.5) | 1275 (0.9) |
| High-risk medical condition |  |  |  |
| Yes | 2095 (32.4) | 2070 (32.1) | 4165 (32.2) |
| No | 4377 (67.6) | 4380 (67.9) | 8759 (67.8) |

\*Two participants received a vaccine at V1 but whether they received the vaccine or the placebo is unknown. Therefore there is a difference of 2 participants in the total number of participants of the SafAS.
†One of the 2 participants who had missing information about the vaccine/placebo was American Indian or Alaska Native. For the other participant, the race was unknown although the ethnicity was Hispanic or Latino.

In both treatment groups, the longest duration of follow-up was 148 days (median 85 days) PD1 and 118 days (median 58 days) PD2 (**Supplementary Appendix Sections 2.4 and 2.5**). The proportion of patients with ≥2 months’ follow-up at the data cut-off date was 67.4% (8,706/12,924) PD1 and 47.2% (5,453/11,543) PD2. Variant distribution according to time and country is shown in **Figure 2**.

**Figure 2:**
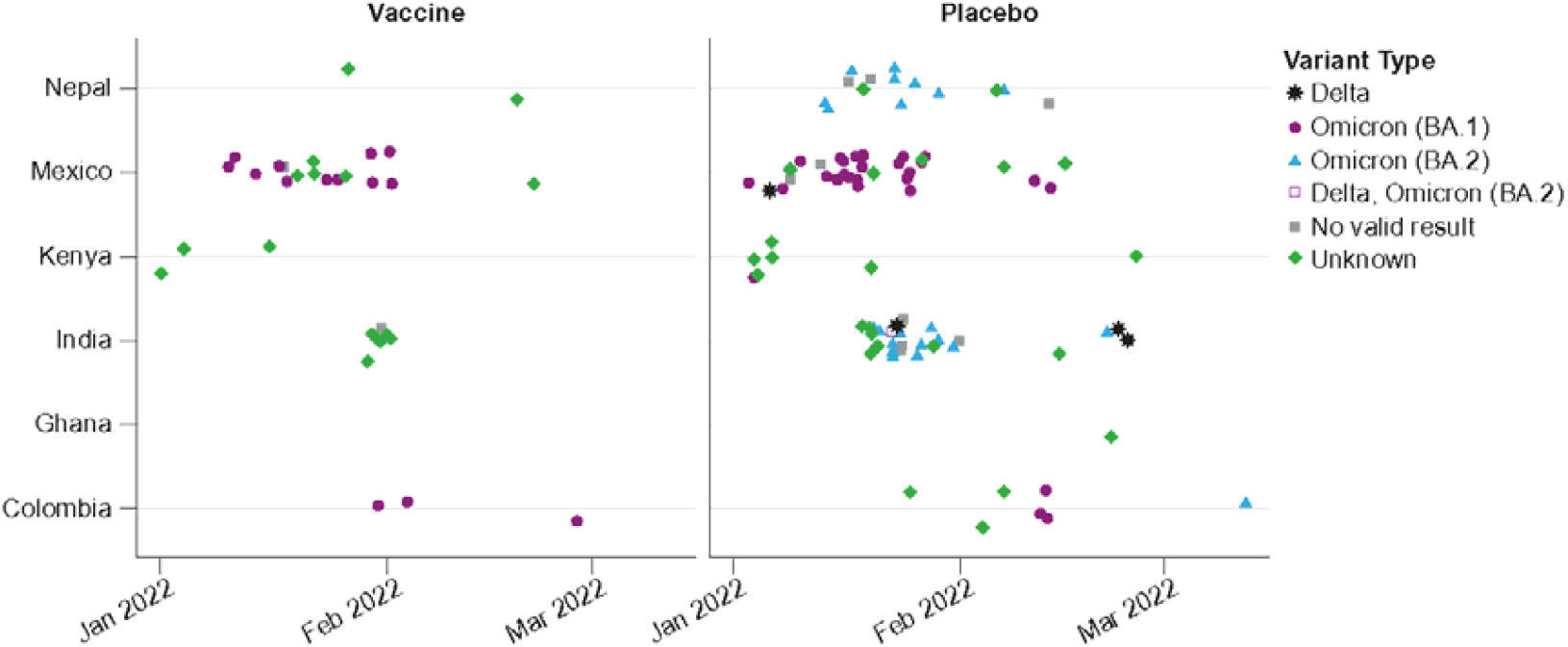
Variant distribution by country and calendar time in all participants, regardless of prior SARS-CoV-2 infections (mFAS-PD2)

### Efficacy

The mFAS-PD2 set comprised 11,416 participants (5736 [50.2%] in the vaccine group; 5680 [49.8%] in the placebo group). 121 symptomatic COVID-19 episodes were reported ≥14 days PD2 (32 in the vaccine vs 89 in the placebo group), with an overall VE of 64.7% (95% CI 46.6; 77.2%) which met the primary efficacy endpoint (**Figure 3**). Similar results were reported in the sensitivity analysis including Ukrainian participants (**Supplementary Appendix Section 2.6**). The cumulative incidence rate of symptomatic COVID-19 was higher in the placebo group than in the vaccine group starting from 14 days after the second dose (**Figure 4**).

**Figure 3:**
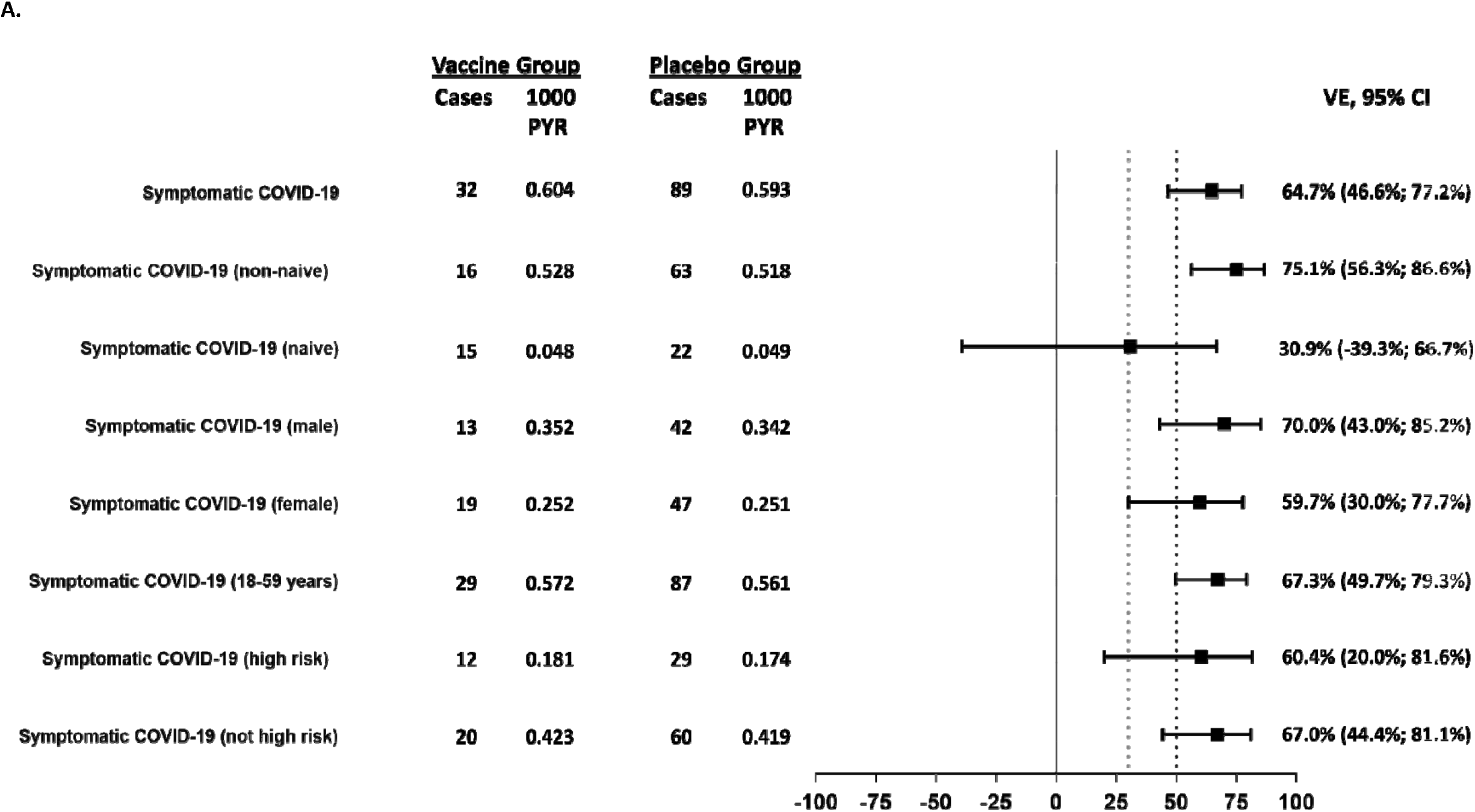
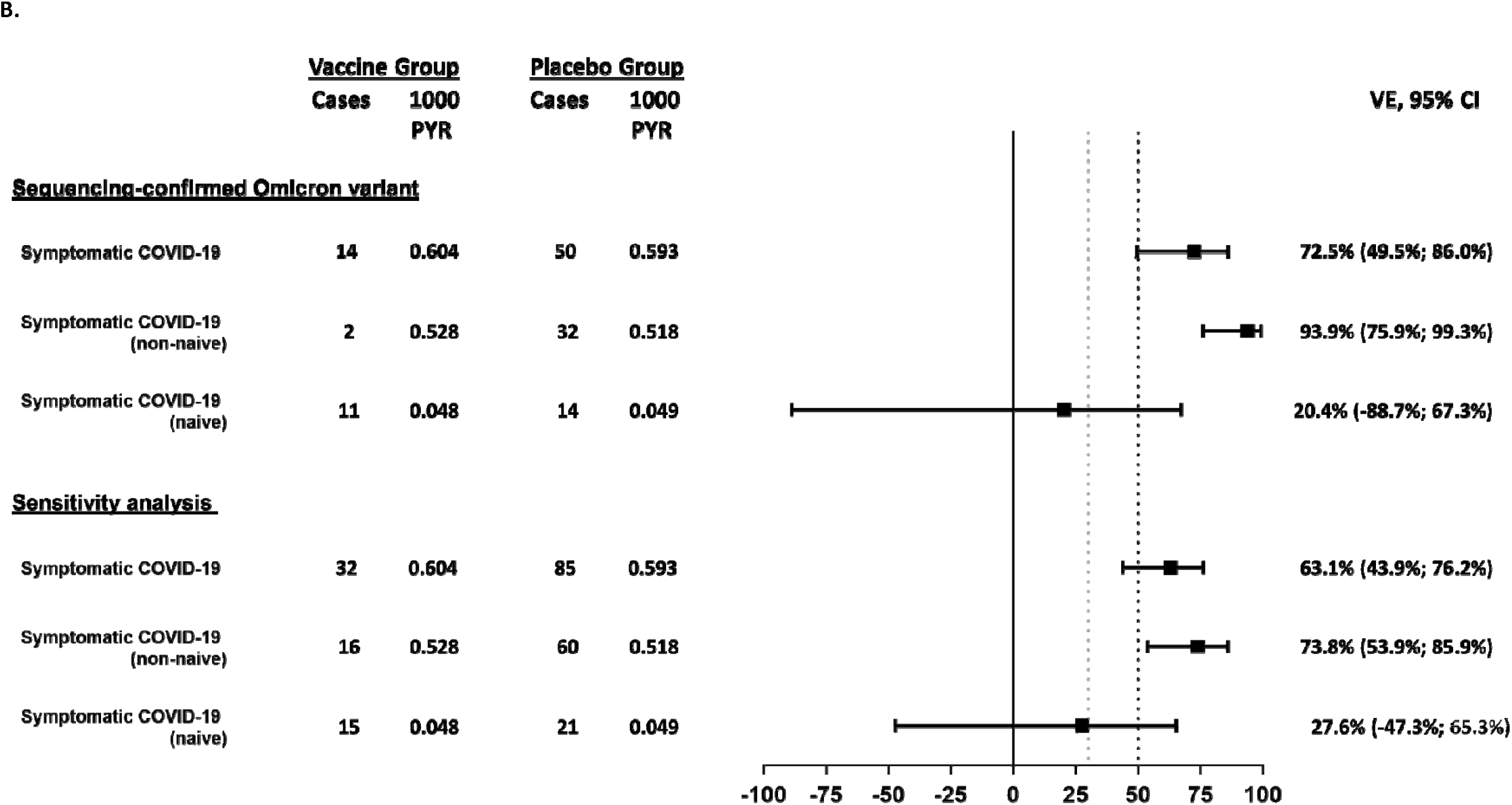
Forest plots for efficacy outcomes against symptomatic disease in all participants and subgroups caused by (A) all variants and (B) for the Omicron variant A. Efficacy outcomes overall and by subgroups for the mFAS-PD2 analysis subset. The success criteria for demonstration of efficacy was defined as a point estimate >50% (black dotted line) and a lower bound confidence interval >30% (grey dotted line). Outcomes with too few cases to reliably calculate vaccine efficacy (severe COVID-19, moderate or worse COVID-19, hospitalization, and symptomatic COVID-19 in participants aged ≥60 years) are not shown. B. Vaccine efficacy is shown for all sequence-confirmed Omicron cases and for the sensitivity analysis, which included sequence confirmed cases and cases for which there were no sequencing results, assuming that the latter group were caused by the Omicron variant as this was the variant that was responsible for most of the symptomatic COVID-19 cases at the time of the study. The success criteria for demonstration of efficacy was defined as a point estimate >50% (black dotted line) and a lower bound confidence interval >30% (grey dotted line). Owing to the low number of cases due to the Delta variant, these are not shown in the Forest plot.

**Figure 4:**
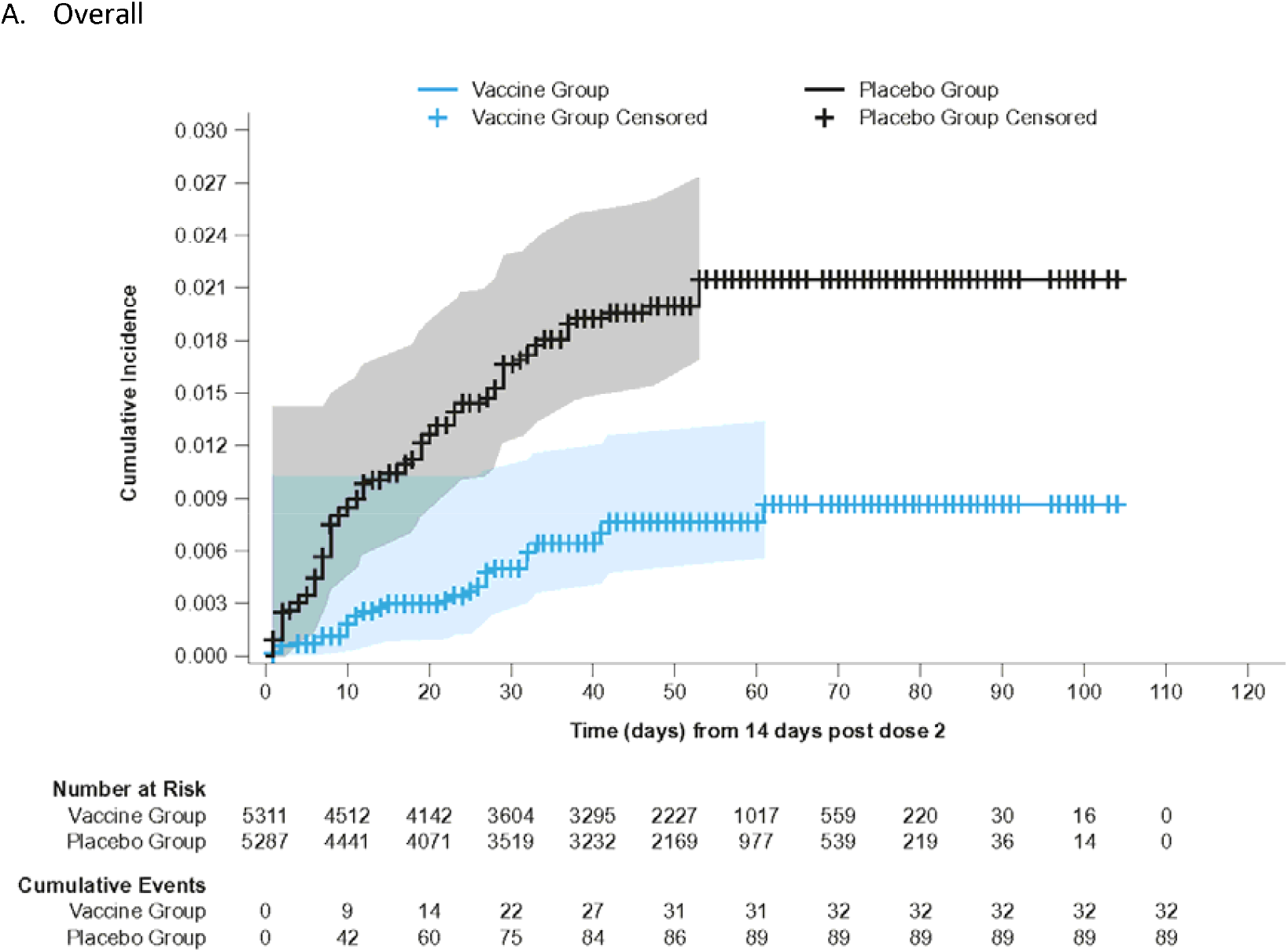
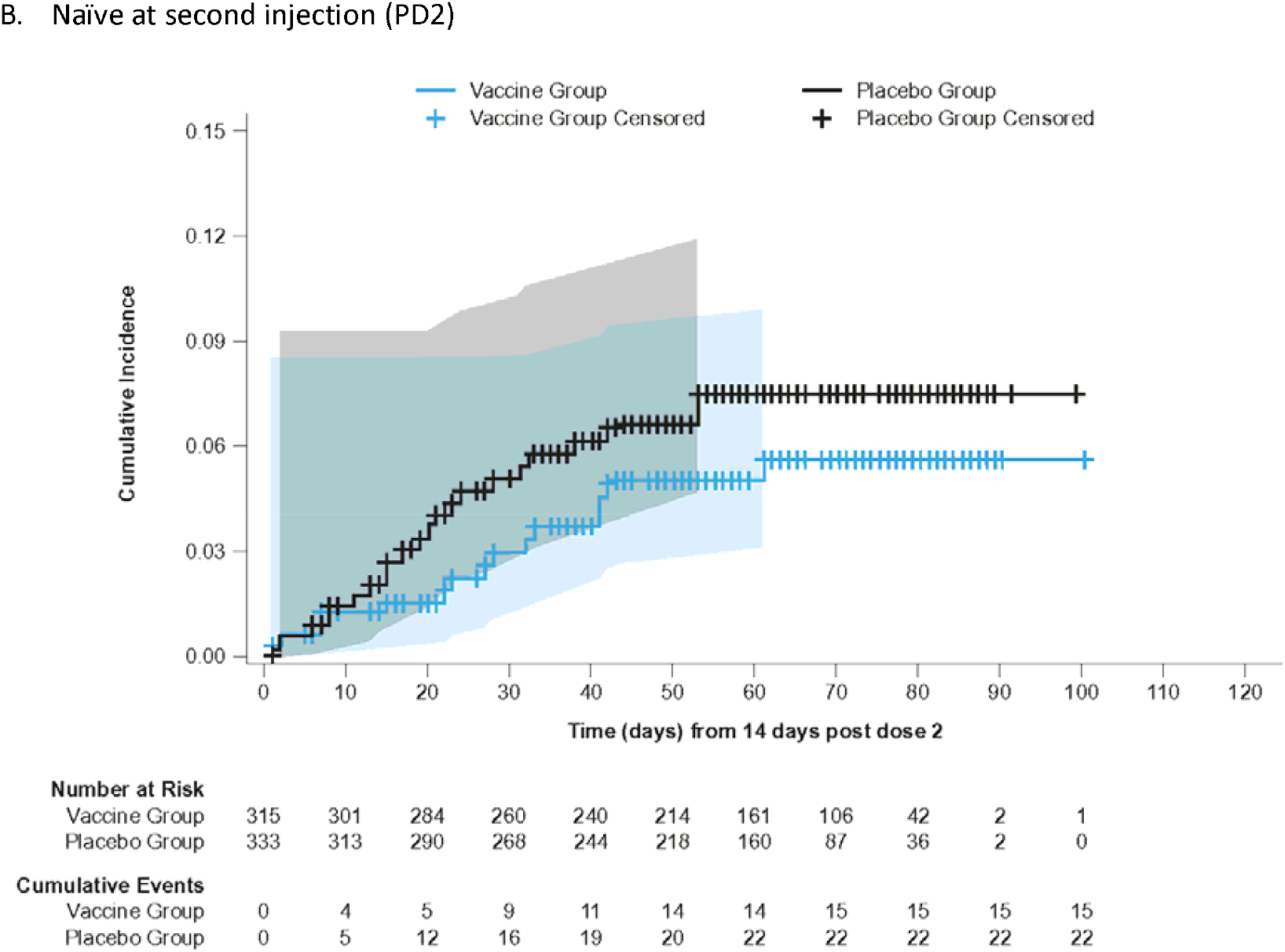
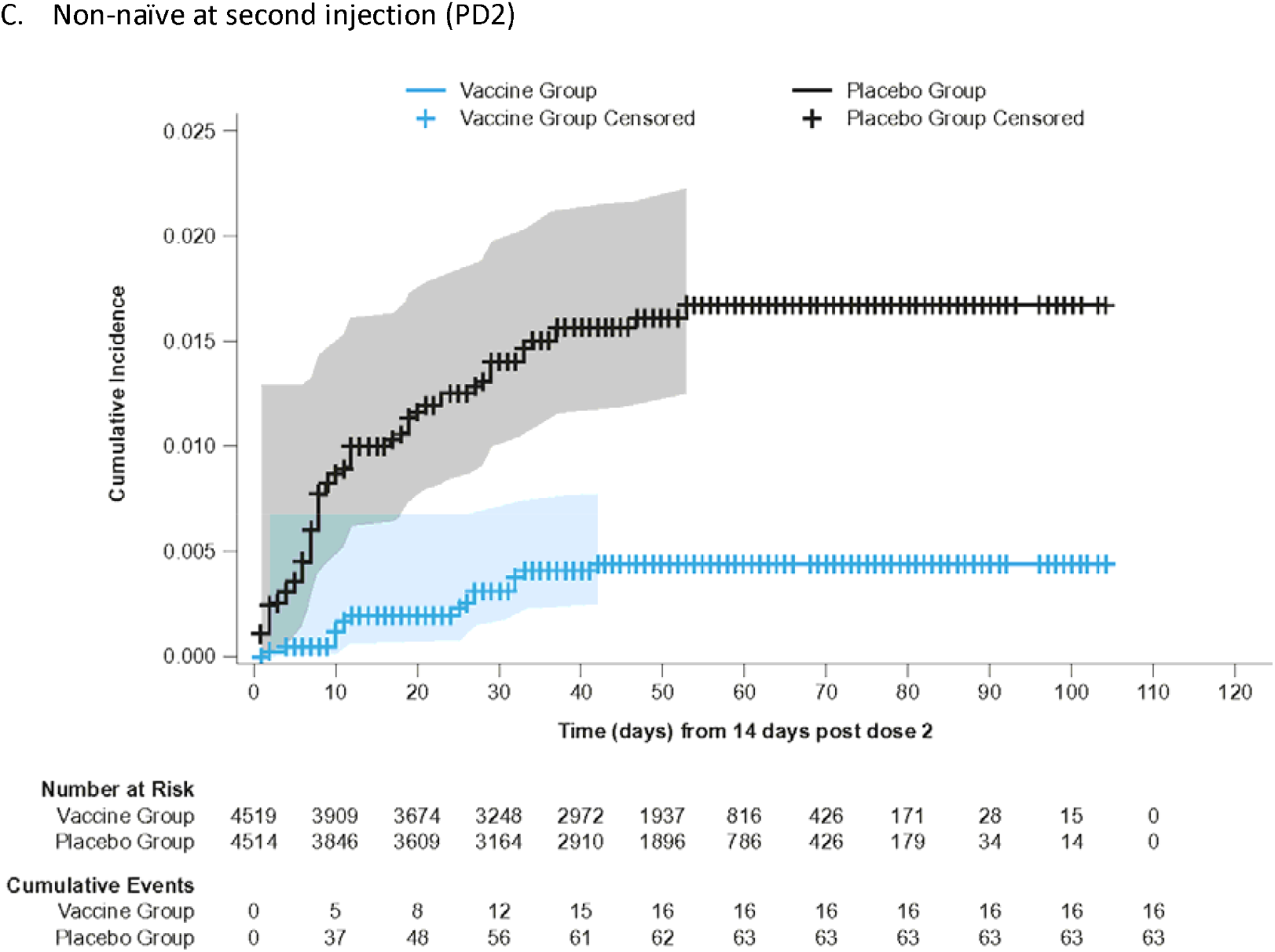
Kaplan-Meier cumulative incidence of symptomatic COVID-19 in the mFAS-PD2 population (overall, naïve and non-naïve populations)

Five participants (three vaccine recipients, two placebo recipients) reported severe COVID-19, and 12 participants reported moderate or worse symptomatic COVID-19 (five vaccine recipients, seven placebo recipients) occurring from 14 days PD2 in mFAS-PD2 participants. Two placebo recipients in the mFAS-PD2 were hospitalized with COVID-19, and there were no deaths associated with COVID-19 reported in the study.

VE against symptomatic COVID-19 infection in non-naïve participants was 75.1% (95% CI: 56.3%; 86.6%), while in naïve participants the point estimate for VE was 30.9% (95% CI -39.3%; 66.7%) (**Figure 3)**. The cumulative incidence was higher in the placebo group than in the vaccine group starting from 14 days PD2 in non-naïve participants and after approximately 30 days PD2 in naïve participants **(Figure 4)**. The overall VE against symptomatic COVID-19 was 60.3% (95% CI 47.1%; 70.5%) PD1 (**Supplementary Appendix Section 2.7**). The higher cumulative incidence in the placebo group started within 14 days PD1 in naïve, non-naïve and all participants in the mFAS-PD1 population (**Supplementary Appendix 2.8**).

Efficacy results against symptomatic disease in all participants and subgroups are shown in **Figure 3** and **Supplementary Appendix Section 2.9**. Efficacy against asymptomatic SARS-CoV-2 infection (assessed in naïve participants only) was 1.2 (95% CI -31.0; 25.5) with 100 cases in the vaccine group and 107 cases in the placebo group (**Supplementary Appendix Section 2.10)**.

### Viral variants

Of the 121 adjudicated cases, the causative viral strain was sequenced in 68 cases (56%), with the majority (63/68) corresponding to the BA.1 and BA.2 subvariants of Omicron and the others corresponding to Delta (4/68). One participant had mixed infection with the Omicron and Delta variants and was included in the analysis for both variants. Results for the other 53 adjudicated cases (approximately 44%) were not available for different reasons (**Supplementary Appendix Section 2.11**).

Among the 68 sequenced cases, 64 were Omicron (14 in the vaccine recipients and 50 in the placebo recipients), with the Omicron-specific VE estimated as 72.5% (95% CI: 49.5; 86.0) (**Figure 3**). Kaplan-Meier analyses showed higher cumulative incidence in the placebo group compared with the vaccine group 14 days PD2 (**Supplementary Appendix Section 2.12**). There was also a favorable case split relating to the Delta variant: no Delta-related COVID-19 cases in the vaccine group versus five cases in the placebo group.

The VE against symptomatic COVID-19 caused by the Omicron or undefined variants (sensitivity analyses) was 63.1% (95% CI 43.9; 76.2%) in all participants, 73.8% (95% CI 53.9; 85.9) in non-naïve participants and 27.6% (95% CI -47.3; 65.3) in naïve participants (**Supplementary Appendix Section 2.13**).

### Safety

A summary of safety outcomes in patients who received at least one injection of vaccine or placebo (SafAS population) are reported in **Table 2** and **Supplementary Appendix Sections 2.14** and **2.15**.

**Table 2:**
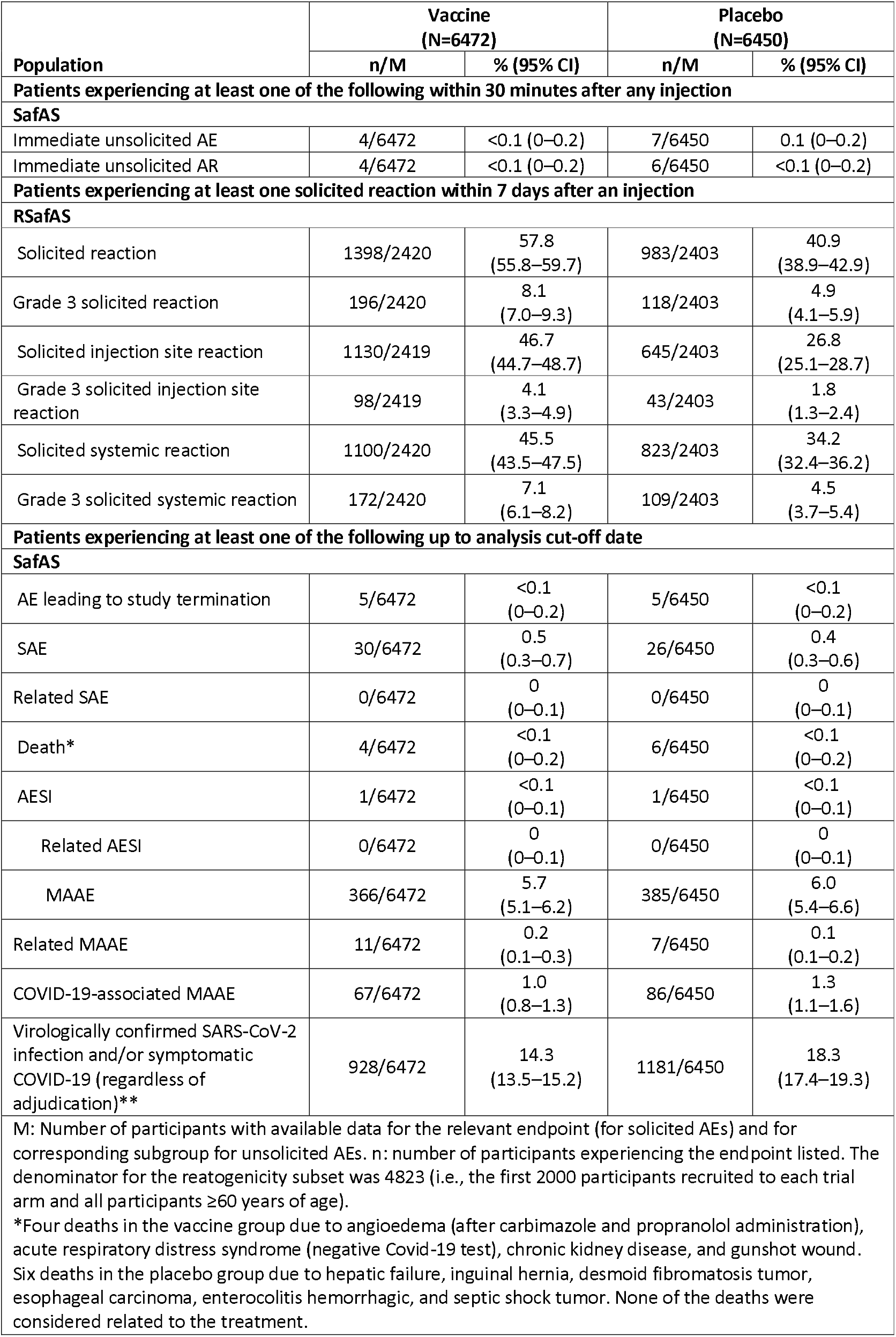
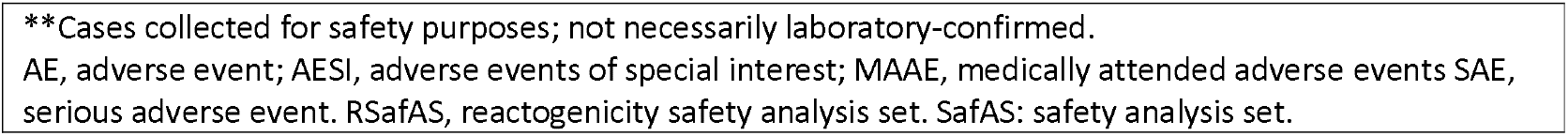
Summary of safety outcomes in patients who received at least one injection (SafAS)

| Population | Vaccine<br>(N=6472) |  | Placebo<br>(N=6450) |  |
| --- | --- | --- | --- | --- |
|  | n/M | % (95% CI) | n/M | % (95% CI) |
| **Patients experiencing at least one of the following within 30 minutes after any injection** |  |  |  |  |
| **SafAS** |  |  |  |  |
| Immediate unsolicited AE | 4/6472 | <0.1 (0–0.2) | 7/6450 | 0.1 (0–0.2) |
| Immediate unsolicited AR | 4/6472 | <0.1 (0–0.2) | 6/6450 | <0.1 (0–0.2) |
| **Patients experiencing at least one solicited reaction within 7 days after an injection** |  |  |  |  |
| **RSafAS** |  |  |  |  |
| Solicited reaction | 1398/2420 | 57.8<br>(55.8–59.7) | 983/2403 | 40.9<br>(38.9–42.9) |
| Grade 3 solicited reaction | 196/2420 | 8.1<br>(7.0–9.3) | 118/2403 | 4.9<br>(4.1–5.9) |
| Solicited injection site reaction | 1130/2419 | 46.7<br>(44.7–48.7) | 645/2403 | 26.8<br>(25.1–28.7) |
| Grade 3 solicited injection site reaction | 98/2419 | 4.1<br>(3.3–4.9) | 43/2403 | 1.8<br>(1.3–2.4) |
| Solicited systemic reaction | 1100/2420 | 45.5<br>(43.5–47.5) | 823/2403 | 34.2<br>(32.4–36.2) |
| Grade 3 solicited systemic reaction | 172/2420 | 7.1<br>(6.1–8.2) | 109/2403 | 4.5<br>(3.7–5.4) |
| **Patients experiencing at least one of the following up to analysis cut-off date** |  |  |  |  |
| **SafAS** |  |  |  |  |
| AE leading to study termination | 5/6472 | <0.1<br>(0–0.2) | 5/6450 | <0.1<br>(0–0.2) |
| SAE | 30/6472 | 0.5<br>(0.3–0.7) | 26/6450 | 0.4<br>(0.3–0.6) |
| Related SAE | 0/6472 | 0<br>(0–0.1) | 0/6450 | 0<br>(0–0.1) |
| Death* | 4/6472 | <0.1<br>(0–0.2) | 6/6450 | <0.1<br>(0–0.2) |
| AESI | 1/6472 | <0.1<br>(0–0.1) | 1/6450 | <0.1<br>(0–0.1) |
| Related AESI | 0/6472 | 0<br>(0–0.1) | 0/6450 | 0<br>(0–0.1) |
| MAAE | 366/6472 | 5.7<br>(5.1–6.2) | 385/6450 | 6.0<br>(5.4–6.6) |
| Related MAAE | 11/6472 | 0.2<br>(0.1–0.3) | 7/6450 | 0.1<br>(0.1–0.2) |
| COVID-19-associated MAAE | 67/6472 | 1.0<br>(0.8–1.3) | 86/6450 | 1.3<br>(1.1–1.6) |
| Virologically confirmed SARS-CoV-2 infection and/or symptomatic COVID-19 (regardless of adjudication)** | 928/6472 | 14.3<br>(13.5–15.2) | 1181/6450 | 18.3<br>(17.4–19.3) |
| M: Number of participants with available data for the relevant endpoint (for solicited AEs) and for corresponding subgroup for unsolicited AEs. n: number of participants experiencing the endpoint listed. The denominator for the reatogenicity subset was 4823 (i.e., the first 2000 participants recruited to each trial arm and all participants ≥60 years of age). |  |  |  |  |
| *Four deaths in the vaccine group due to angioedema (after carbimazole and propranolol administration), acute respiratory distress syndrome (negative Covid-19 test), chronic kidney disease, and gunshot wound. Six deaths in the placebo group due to hepatic failure, inguinal hernia, desmoid fibromatosis tumor, esophageal carcinoma, enterocolitis hemorrhagic, and septic shock tumor. None of the deaths were considered related to the treatment. |  |  |  |  |

\*\*Cases collected for safety purposes; not necessarily laboratory-confirmed. AE, adverse event; AESI, adverse events of special interest; MAAE, medically attended adverse events SAE, serious adverse event. RSafAS, reactogenicity safety analysis set. SafAS: safety analysis set.

For both the vaccine and placebo groups, immediate unsolicited AEs and adverse reactions (ARs) ≤30 minutes after any injection were reported by <0.1%. In the reactogenicity subset (N=4,823), solicited reactions (SISRs and SSRs) ≤7 days after any injection occurred in 57.8% vaccine recipients and 40.9% placebo recipients (**Figure 5**).

**Figure 5:**
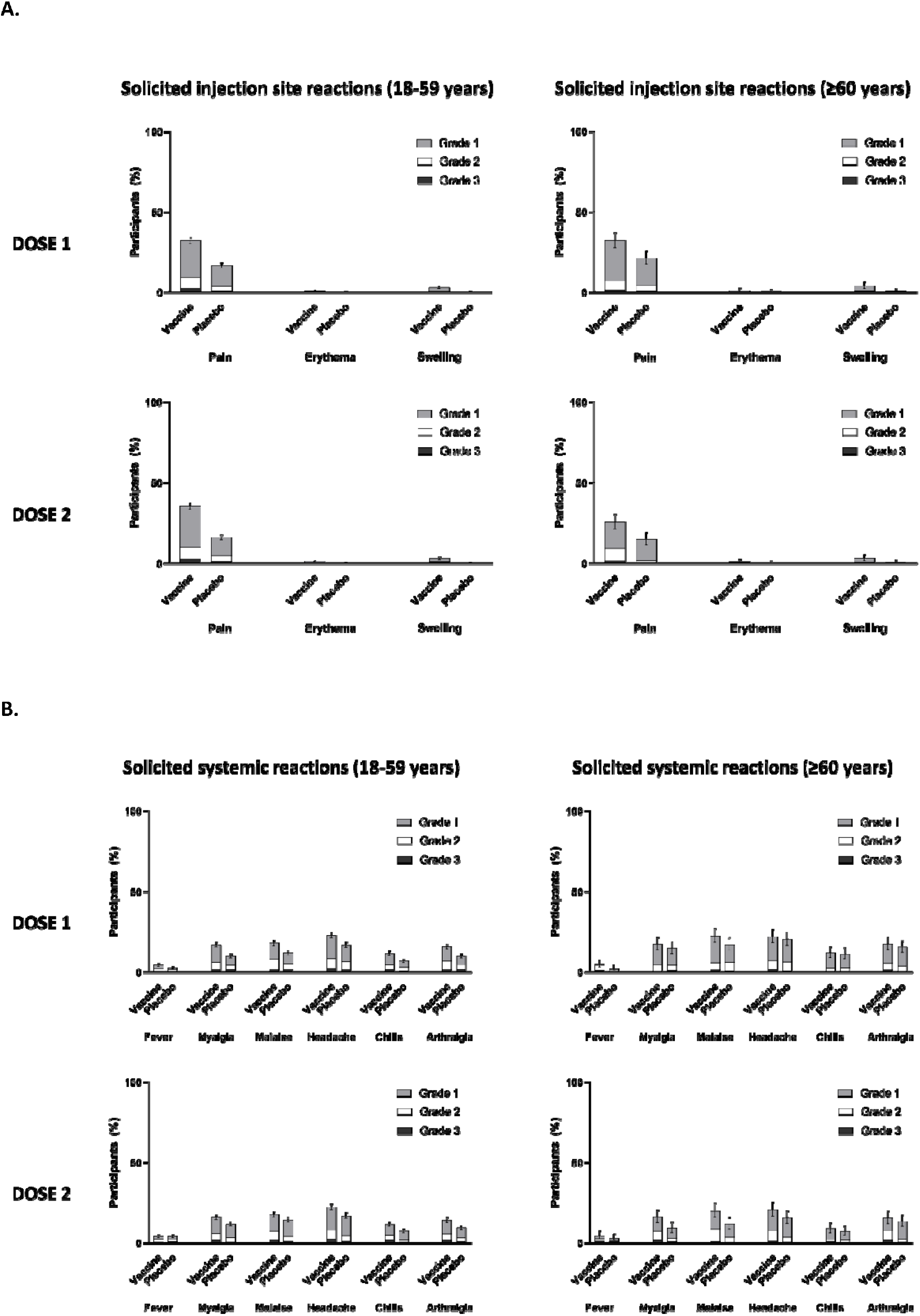
(A) Proportion of participants with solicited injection site reactions within 7 days of each study injection in participants aged 18–59 years and participants aged ≥60 years; (B) the proportion of participants with solicited systemic reactions within 7 days of each study injection in participants aged 18–59 years and participants aged ≥60 years

Grade 3 solicited reactions were reported by 8.1% of vaccine recipients and 4.9% of the placebo recipients within 7 days after any injection, with comparable frequency PD1 and PD2 in the vaccine group (**Table 2; Figure 5; Supplementary Appendix Section 2.14**).

The proportion of MAAEs reported was similar in the vaccine (5.7%) and placebo (6.0%) groups. The proportion of AESIs, SAEs and deaths were <1% in both study arms; no AE, AESI, SAE or death was deemed to be treatment related. There were no reported cases of thrombosis with thrombocytopenia syndrome, myocarditis, pericarditis, Bell’s Palsy, or Guillain–Barré syndrome.

## Discussion

This is the first report of an efficacy trial conducted with a variant COVID-19 vaccine. In this Phase 3 study evaluating a bivalent vaccine as a primary series during the period of predominant Omicron (BA.1 and BA.2) circulation, the primary objective of demonstrating efficacy against symptomatic COVID-19 of >50%, with a lower bound of the 95% confidence interval >30%, in all participants was met.

The epidemiological context for this efficacy trial is markedly different from those conducted at the pandemic’s onset.^18,19^ A large proportion of participants had serological evidence of previous infection representative of the epidemiological situation at the time of the study. Thus, the VE against symptomatic COVID-19 in non-naïve participants of 75% observed in this study starting 14 days PD1 is of particular relevance. This also suggests the potential use of the vaccine as a booster dose at this stage of the pandemic when most of the population have already been exposed to the virus or have been vaccinated. Lower VE was observed in naïve individuals, albeit the number of participants in this sub-group was limited. These are consistent with observations in other efficacy trials^20,21^ and the high antibody titres observed in animal studies.^11^

During the surveillance period, two major variants were circulating: Omicron (BA.1 and BA.2 subvariants) and to a lesser extent Delta, with no cases of BA.4 and BA.5. Thus, the data reported here is the first assessment of clinical efficacy of a COVID-19 vaccine against the Omicron variant. Since sequencing results were unavailable in approximately 44% of the cases in the mFAS-PD2, we conducted sensitivity analyses that assumed these cases were caused by Omicron variants, and VE was also demonstrated.

Primary immunization with two doses of prototype vaccines provided limited protection against symptomatic disease caused by the Omicron variant. We demonstrated efficacy against Omicron with two doses of a Beta-containing variant as opposed to previous reports of high efficacy against Omicron following three doses of mRNA vaccines.^22^ A BNT162b2 or mRNA-1273 booster after a primary course substantially increased protection, but that protection waned over time.^23^ Variant-updated COVID-19 vaccines^24^ and booster vaccines incorporating Omicron subvariants are under development or are authorized for use. Their use has been endorsed by global regulators provided that novel COVID-19 booster vaccines containing alternative variants still confer adequate protection against Omicron and other VOCs. Our Beta strain-containing vaccine confers protection against newly emergent variants, thus providing clinical evidence that broad cross-protection can be conferred by such vaccines, and challenges the current paradigm of variant-chasing vaccine strain composition.

While the exact mechanism of cross-protection is unknown, it may be primarily related to the B1.3.5.1 component of the bivalent vaccine. Substitutions in the Beta variant spike at positions K417N, E484K, N501Y may provide new antibody epitopes which are well-positioned to provide cross-neutralizing immunogenicity against a wide array of variants including contemporary circulating strains.^15^ The results of this study in Omicron-confirmed cases suggests the potential for a Beta variant containing variant vaccine to be used as a part of a booster program, and a beta variant containing vaccine (VidPrevtyn Beta) has now been recommended as a booster in adults previously vaccinated with a mRNA or adenoviral vector COVID-19 vaccine.^25^ Results from a booster study in individuals previously primed with the CoV2 preS dTM-AS03 (D614) vaccine or with other approved mRNA and adenovirus-vectored vaccines, confirmed that a booster with an CoV2 preS dTM-AS03 (B.1.351, Beta) vaccine delivered an immune response comparable to that of the bivalent (ancestral + Beta variant) booster (in press).

The number of severe COVID-19 cases or hospitalizations was limited; however, all hospitalized cases were observed in the placebo group. The few severe and hospitalized cases may have been due to the Omicron variant leading to milder COVID-19 disease versus other variants, particularly as most participants had already experienced a prior SARS-CoV-2 infection.^26^ Additionally, most participants in this study were younger adults aged 18–59 years with lower risk of severe COVID-19 than older people.^27,28^ Of note, the VE in participants aged 18–59 years with risk factors for severe COVID 19 was similar to that in the same age group without risk factors.

The bivalent vaccine showed an acceptable reactogenicity profile in this study; after both doses, AEs were mostly mild to moderate and transient, regardless of participant age or prior infection. Injection-site and systemic reactions were each reported by less than half of participants in the reactogenicity subset. These rates may indicate potentially less reactogenicity compared with mRNA-based licensed vaccines, although these vaccines have not been evaluated together in the context of a single trial.^29,30^ No cases of myocarditis, pericarditis or thrombosis with thrombocytopenia syndrome were reported during the observed 2–3 months of safety follow-up, which have previously been reported after vaccination with other vaccines.^31-41^

Our study has limitations. Due to the limited number of older adults (≥60 years) enrolled in the trial, VE could not be accurately estimated in this age group. This was most likely due to the roll-out of vaccines authorized for emergency use in this age category available at the time of the study. The limited number of hospitalized and severe cases prevented any conclusions for VE against these outcomes. The short duration of follow-up (median length of follow up PD2 was 58 days) also precluded conclusions on the durability of the vaccine’s protection and long-term safety. Because immunogenicity results were not available, correlates of protection could not be assessed. While sequencing was attempted on all primary endpoint cases, results were only available in approximately 56% of primary endpoints. We observed a higher rate of missing sequence data in the vaccine group (56%) compared to the placebo group (39%). One explanation for this observation is the potential impact of the vaccine on reducing viral load. Although the higher rate of missing data in the vaccine group may bias the variant-specific efficacy estimates, sensitivity analyses confirmed efficacy against Omicron.

## Conclusions

Our results demonstrate the clinical efficacy of a beta variant containing vaccine to protect against different SARS-CoV-2 variants, including Omicron (BA.1 and BA.2), and an acceptable safety profile in adults <60 years old. These data show that vaccines developed with an antigen from a non-predominant strain can confer cross-protection against newly emergent variants.

## Data Availability

Qualified researchers can request access to patient-level data and related study documents, including the clinical study report, study protocol with any amendments, blank case report forms, statistical analysis plan, and dataset specifications. Patient-level data will be anonymized and study documents will be redacted to protect the privacy of trial participants. Further details on Sanofi's data sharing criteria, eligible studies, and process for requesting access can be found at https://vivli.org/.

## Acknowledgements

The authors thank all participants, investigators, and study site personnel who took part in this study, including: those from the Ghana Health Service Kintampo Health Research Centre; Kwame Ayisi Boateng, Anthony Afum-Adjei Awuah; Melvin Agbogbatey; Ebenezer Ahenkan, and Esther Daley Matey from the Kwame Nkrumah University of Science and Technology (KNUST), Kumasi, Ghana; Claudia Pimentel from the Instituto Nacional de Pediatría, México; Victor Mudhune Otieno, Grace Mugure Mboya, Zipporah Nyamoita Buko, and Taraz Samandari from the KEMRI CGHR Kisumu Kenya; Andrea Accini Valencia, Melissa Accini Valencia, Andrea de Moya, and Yineth Conrado, from the IPS Cenatro Cientifico S.A.A, Colombia; Kenneth K Ngure, Eddah Mbugua, Stanley Ndwiga, Stephen Maina Gakuo, Philip Mwangi, David Chege, Jacinta Nyokabi, and Mercy Nyawira, from the PHRD-Thika KEMRI Clinic, Thika, Kenya; Nichlous Ssebudde, Mary Grace Nalubega, Jenifer Alaba, Emmanuel Obonyo, Raymondo Oola, Harriet Tino, Geoffrey Magombe, Vanon Kyehayo, Ritah Norah Nalybwama, Rebecca Asiimire, Denis Wokorac, Agnes Alimo, and Abigail Link, from the SICRA site at Lira Regional Referral Hospital, Uganda; Korutaro Violet, Elyanu Peter James, Baguma Allan, Kekitiinwa Adeodata, Sekabira Rogers, Ssebunnya Billy, and Ashaba Justus from the Baylor College of Medicine Children’s Foundation-Uganda; Maricianah Onono, Imeldah Wakhungu, Kevin Onyango, Dismas Congo, Samya Rashid, Florence Ondiek, George Otieno, Job Ouma, Donnavane Ondego, Maqline Juma, Penina Amboka, Perez Odhiambo, Mildred Obare, Teresia Otieno, Caren Awinja, and Lizzie Kabete, from the Kenya Medical Research Institute – Centre for Microbiological Research, Research Care and Training Program; Lorena Buitrago from the Centro de Atencion e Investigacion Medica S.A.S. – Caimed S.A.S – sede Bogotá; Patrick Ansah, Oscar Bangre, Michael Bandasua Kaburise and Francis Broni, from the Navrongo Health Research Centre, Ghana; Shelly Ramirez, Gail Broder, Liz Breisemeister, Jim Kublin, David Benkeser, and John Hural, from the Fred Hutch Cancer Center, Seattle, WA, USA; Seyram Kaali, Samuel Harrison, Prince Agyapong, Felicia Serwah, Cynthia Bema, Elvis Eilson, Afia Korkor Opare Yeboah, Dennis Adu-Gyasi, Elisha Adeniji, Owusu Boahen, and Zakariah Buwah from the Research and Development Division, Ghana Health Service, Kintampo North Municipality, Kintampo; Oumou Maiga-Ascofare from the Kumasi Center for Collaborative Research in Tropical Medicine, Kumasi. The authors would also like to thank the Ministry of Health & Population, Government of Nepal for providing swift approval to conduct this trial in Nepal The authors acknowledge Steven Goodrick PhD and Nicola Truss PhD of inScience Communications, Springer Healthcare Ltd, London, UK, for providing medical writing assistance with the preparation of this manuscript, funded by Sanofi. The authors also thank Hanson Geevarghese for providing editorial assistance and manuscript coordination on behalf of Sanofi.

Funding was provided by Sanofi and by federal funds from the Biomedical Advanced Research and Development Authority, part of the office of the Administration for Strategic Preparedness and Response at the U.S. Department of Health and Human Services under contract number HHSO100201600005I, and in collaboration with the U.S. Department of Defense Joint Program Executive Office for Chemical, Biological, Radiological and Nuclear Defense under contract number W15QKN-16-9-1002. The views presented here are those of the authors and do not purport to represent those of the Department of the Army. This work was done in collaboration with GSK, who provided access to, and use of, the AS03 Adjuvant System.

The NIAID provides grant funding to the HIV Vaccine Trials Network (HVTN) Leadership and Operations Center (UM1 AI 68614HVTN), the Statistics and Data Management Center (UM1 AI 68635), the HVTN Laboratory Center (UM1 AI 68618), the HIV Prevention Trials Network Leadership and Operations Center (UM1 AI 68619), the AIDS Clinical Trials Group Leadership and Operations Center (UM1 AI 68636), and the Infectious Diseases Clinical Research Consortium leadership group 5 (UM1 AI 148684-03).

## References

1. Cai Y, Zhang J, Xiao T, et al. Structural basis for enhanced infectivity and immune evasion of SARS-CoV-2 variants. Science 2021;373:642–8.

2. Emary KRW, Golubchik T, Aley PK, et al. Efficacy of ChAdOx1 nCoV-19 (AZD1222) vaccine against SARS-CoV-2 variant of concern 202012/01 (B.1.1.7): an exploratory analysis of a randomised controlled trial. Lancet 2021;397:1351–62.

3. Madhi SA, Baillie V, Cutland CL, et al. Efficacy of the ChAdOx1 nCoV-19 Covid-19 Vaccine against the B.1.351 Variant. N Engl J Med 2021;384:1885–98.

4. Shinde V, Bhikha S, Hoosain Z, et al. Efficacy of NVX-CoV2373 Covid-19 Vaccine against the B.1.351 Variant. N Engl J Med 2021;384:1899–909.

5. Desai D, Khan AR, Soneja M, et al. Effectiveness of an inactivated virus-based SARS-CoV-2 vaccine, BBV152, in India: a test-negative, case-control study. The Lancet Infectious Diseases.

6. Buchan SA, Chung H, Brown KA, et al. Effectiveness of COVID-19 vaccines against Omicron or Delta symptomatic infection and severe outcomes. medRxiv 2022:2021.12.30.21268565.

7. Collie S, Champion J, Moultrie H, Bekker L-G, Gray G. Effectiveness of BNT162b2 Vaccine against Omicron Variant in South Africa. New England Journal of Medicine 2021;386:494–6.

8. Adapted vaccine targeting BA.4 and BA.5 Omicron variants and original SARS-CoV-2 recommended for approval European Medicines Agency, 2022. (Accessed 4 Oct, 2022, at https://www.ema.europa.eu/en/news/adapted-vaccine-targeting-ba4-ba5-omicron-variants-original-sars-cov-2-recommended-approval.)

9. Coronavirus (COVID-19) Update: FDA Authorizes Moderna, Pfizer-BioNTech Bivalent COVID-19 Vaccines for Use as a Booster Dose. US Food and Drug Administration, 2022. (Accessed 4 Oct, 2022, at https://www.fda.gov/news-events/press-announcements/coronavirus-covid-19-update-fda-authorizes-moderna-pfizer-biontech-bivalent-covid-19-vaccines-use.)

10. Berry CP V; Anosova, N; Kishko, M; Huang, D; Tibbitts, T; Raillard, A; Gautheron, S; Cummings, S; Bangari, D; Kar, S; Atyeo, C; Deng, Y; Alter, G; Gutzeit, C; Koutsoukos, M; Chicz, R; Lecouturier, V. A Beta-containing bivalent SARS-CoV-2 spike protein vaccine candidate with AS03 elicits durable and broad neutralization of variants including Omicron in macaques and confers protection in hamsters. Preprint 2022.

11. Pavot V, Berry C, Kishko M, et al. Protein-based SARS-CoV-2 spike vaccine booster increases cross-neutralization against SARS-CoV-2 variants of concern in non-human primates. Nature communications 2022;13:1699.

12. Joffe S, Babiker A, Ellenberg SS, et al. Data and Safety Monitoring of COVID-19 Vaccine Clinical Trials. The Journal of infectious diseases 2021;224:1995–2000.

13. Garçon N, Vaughn DW, Didierlaurent AM. Development and evaluation of AS03, an Adjuvant System containing α-tocopherol and squalene in an oil-in-water emulsion. Expert review of vaccines 2012;11:349–66.

14. Goepfert PA, Fu B, Chabanon AL, et al. Safety and immunogenicity of SARS-CoV-2 recombinant protein vaccine formulations in healthy adults: interim results of a randomised, placebo-controlled, phase 1-2, dose-ranging study. Lancet Infect Dis 2021;21:1257–70.

15. Sridhar S, Joaquin A, Bonaparte MI, et al. Safety and immunogenicity of an AS03-adjuvanted SARS-CoV-2 recombinant protein vaccine (CoV2 preS dTM) in healthy adults: interim findings from a phase 2, randomised, dose-finding, multicentre study. Lancet Infect Dis 2022.

16. Shrestha L, Lin MJ, Xie H, et al. Clinical Performance Characteristics of the Swift Normalase Amplicon Panel for Sensitive Recovery of Severe Acute Respiratory Syndrome Coronavirus 2 Genomes. The Journal of molecular diagnostics : JMD 2022;24:963–76.

17. Covid_Swift _pipeline. 2022. (Accessed 11 November, 2022, at https://github.com/greninger-lab/covid_swift_pipeline.)

18. Baden LR, El Sahly HM, Essink B, et al. Efficacy and Safety of the mRNA-1273 SARS-CoV-2 Vaccine. N Engl J Med 2021;384:403–16.

19. Polack FP, Thomas SJ, Kitchin N, et al. Safety and Efficacy of the BNT162b2 mRNA Covid-19 Vaccine. N Engl J Med 2020;383:2603–15.

20. Hall V, Foulkes S, Insalata F, et al. Protection against SARS-CoV-2 after Covid-19 Vaccination and Previous Infection. N Engl J Med 2022;386:1207–20.

21. Murugesan M, Mathews P, Paul H, Karthik R, Mammen JJ, Rupali P. Protective effect conferred by prior infection and vaccination on COVID-19 in a healthcare worker cohort in South India. PLoS One 2022;17:e0268797.

22. Accorsi EK, Britton A, Fleming-Dutra KE, et al. Association Between 3 Doses of mRNA COVID-19 Vaccine and Symptomatic Infection Caused by the SARS-CoV-2 Omicron and Delta Variants. Jama 2022;327:639–51.

23. Adams K, Rhoads JP, Surie D, et al. Vaccine effectiveness of primary series and booster doses against covid-19 associated hospital admissions in the United States: living test negative design study. Bmj 2022;379:e072065.

24. Interim statement on decision-making considerations for the use of variant updated COVID-19 vaccines.. World Health Organisation, 2022. (Accessed 30 June, 2022, at https://www.who.int/news/item/17-06-2022-interim-statement-on-decision-making-considerations-for-the-use-of-variant-updated-covid-19-vaccines.)

25. EMA recommends approval of VidPrevtyn Beta as a COVID 19 booster vaccine. European Medicines Agency. (Accessed 14 Novemeber, 2022, at https://www.ema.europa.eu/en/news/ema-recommends-approval-vidprevtyn-beta-covid-19-booster-vaccine.)

26. Meo SA, Meo AS, Al-Jassir FF, Klonoff DC. Omicron SARS-CoV-2 new variant: global prevalence and biological and clinical characteristics. European review for medical and pharmacological sciences 2021;25:8012–8.

27. Zhang H, Wu Y, He Y, et al. Age-Related Risk Factors and Complications of Patients With COVID-19: A Population-Based Retrospective Study. Front Med (Lausanne) 2021;8:757459.

28. Levin AT, Hanage WP, Owusu-Boaitey N, Cochran KB, Walsh SP, Meyerowitz-Katz G. Assessing the age specificity of infection fatality rates for COVID-19: systematic review, meta-analysis, and public policy implications. Eur J Epidemiol 2020;35:1123–38.

29. Spikevax: summary of product characteristics. European Medicines Agency, 2022. (Accessed 7 November, 2022, at https://www.ema.europa.eu/en/documents/product-information/spikevax-previously-covid-19-vaccine-moderna-epar-product-information_en.pdf.)

30. Comirnaty: summary of product characteristics. European Medicines Agency, 2022. (Accessed 7 November, 2022, at https://www.ema.europa.eu/en/documents/product-information/comirnaty-epar-product-information_en.pdf.)

31. Lai FTT, Li X, Peng K, et al. Carditis After COVID-19 Vaccination With a Messenger RNA Vaccine and an Inactivated Virus Vaccine : A Case-Control Study. Ann Intern Med 2022;175:362–70.

32. Patone M, Mei XW, Handunnetthi L, et al. Risks of myocarditis, pericarditis, and cardiac arrhythmias associated with COVID-19 vaccination or SARS-CoV-2 infection. Nat Med 2022;28:410–22.

33. Husby A, Hansen JV, Fosbøl E, et al. SARS-CoV-2 vaccination and myocarditis or myopericarditis: population based cohort study. Bmj 2021;375:e068665.

34. Karlstad Ø, Hovi P, Husby A, et al. SARS-CoV-2 Vaccination and Myocarditis in a Nordic Cohort Study of 23 Million Residents. JAMA Cardiol 2022;7:600–12.

35. Mevorach D, Anis E, Cedar N, et al. Myocarditis after BNT162b2 mRNA Vaccine against Covid-19 in Israel. The New England journal of medicine 2021;385:2140–9.

36. Witberg G, Barda N, Hoss S, et al. Myocarditis after Covid-19 Vaccination in a Large Health Care Organization. The New England journal of medicine 2021;385:2132–9.

37. Montgomery J, Ryan M, Engler R, et al. Myocarditis Following Immunization With mRNA COVID-19 Vaccines in Members of the US Military. JAMA Cardiol 2021;6:1202–6.

38. Oster ME, Shay DK, Su JR, et al. Myocarditis Cases Reported After mRNA-Based COVID-19 Vaccination in the US From December 2020 to August 2021. Jama 2022;327:331–40.

39. Husby A, Køber L. COVID-19 mRNA vaccination and myocarditis or pericarditis. The Lancet 2022;399:2168–9.

40. Hafeez MU, Ikram M, Shafiq Z, et al. COVID-19 Vaccine-Associated Thrombosis With Thrombocytopenia Syndrome (TTS): A Systematic Review and Post Hoc Analysis. Clinical and Applied Thrombosis/Hemostasis 2021;27:10760296211048815.

41. Novavax HCP Fact Sheet 09122022. US Food and Drug Administration, 2022. (Accessed 13 October, 2022, at https://www.fda.gov/media/159897/download.)

