## Supplementary Materail for "Efficacy of a bivalent (D614 + B.1.351) SARS-CoV-2 Protein Vaccine" (from another version of this preprint: version 1, 2022-12-06)

### 1.0 Supplementary Methods

#### 1.1 Sites participating in the study

| **Principal Investigator** | **Location** | **City** |
| --- | --- | --- |
| **Colombia** | | |
| Maria Angelica Granados | Centro de Atencion e Investigacion Medica S.A.S. – Caimed S.A.S – sede Bogotá | Bogotá |
| Alberto Cadena Bonfanti | Clinica de la Costa | Barranquilla |
| Hugo Macareno Arroyo | Fundacion Hospital Universidad del Norte | Soledad |
| Eduardo Lopez-Medina | Centro de Estudios en Infectología Pediátrica CEIP, Department of Pediatrics Universidad del Valle and Clinica Imbanaco Grupo Quironsalud | Cali |
| Humberto Reynales | Centro de Atención e Investigación Médica S.A.S. – CAIMED Chía | Chía |
| Jaime Augusto Carrillo | Centro de Atención e Investigación Médica S.A.S.– CAIMED Girardot | Girardot |
| Jose Accini | IPS Centro Cientifico | Barranquilla |
| Hector Velásquez | Centro de Atención e Investigación Médica S.A.S. – sede Acacías | Acacias |
| Edith Rodriguez | Centro de Atención e investigación Médica CAIMED S.A.S | Aguazul |
| Erwin Pardo | Centro de Atencion e investigacion Medica CAIMED SAS | Armenia |
| **Ghana** | | |
| Nana Akosua Ansah | Navrongo Health Research Centre (NHRC) | Navrongo |
| Kwaku Poku Asante | Research and Development Division, Ghana Health Service, Kintampo North Municipality | Kintampo |
| John Humphrey Amuasi | Kwame Nkrumah University of Science and Technology & Kumasi Center for Collaborative Research in Tropical Medicine | Kumasi |
| **India** | | |
| Manish Jain | Maharaja Agrasen Superspeciality Hospital | Jaipur |
| Jinen Shah | Aartham Multi Super Speciality Hospital | Ahmedabad |
| Chandramani Singh | All India Institute of Medical Science (AIIMS) | Patna |
| Jitendra Singh Kushwaha | Prakhar Hospital Private Limited | Kanpur |
| Satyajit Mohapatra | SRM Medical College & Research Centre, | Chennai |
| Amit Suresh Bhate | Jeevan Rekha Hospital | Belagavi |
| Nitin Khandelwal | Vidarbha Institute of Medical Sciences | Nagpur |
| Veer Bahadur Singh | Jawaharlal Nehru Medical College Campus | Ajmer |
| Chandan Das | Institute of Medical Sciences and Sum Hospital | Bhubaneswar |
| **Kenya** | | |
| Videlis Nduba | Kenya Medical Research Institute- Centre for Respiratory Disease Research | Nairobi |
| Lucas Otieno Tina | Kenya Medical Research Institute (KEMRI)/USAMRD-Africa/Kenya | Kisumu |
| Samuel Gurrion Ouma | Kenya Medical Research Institute CGHR, HIVR Division | Kisumu |
| Fredrick Sawe | Kenya Medical Research Institute – US Army Medical Research | Kericho |
| Abraham Siika | Moi University CRC | Eldoret |
| Mansoor Saleh | Aga Khan University Hospital | Nairobi |
| Kishorchandra Mandaliya | Ganjoni Clinic | Mombasa |
| Nelly Mugo | Kenya Medical Research Institute – Partners in Health Research & Development | Thika |
| Janet Oyieko | Kenya Medical Research Institute – Butere County Hospital | Butere |
| Elisabeth Bukusi | Kenya Medical Research Institute – Centre for Microbiological Research, Research Care and Training Program | Kisumu |
| **Mexico** | | |
| Luis Espinoza | Arke SMO SA de CV | Veracruz |
| Maria Otero | Hospital Civil "Fray Antonio Alcalde" | Guadalajara |
| Rafael Rivero | Centro de Investigación Clínica del Pacífico S.A. de C.V. | Acapulco |
| Javier Martínez | Morales Vargas Centro de investigación | León |
| Sandra Villagómez-Martínez | Hospital General de Temixco, en acuerdo con el Instituto Nacional de Pediatría Unidad de Apoyo a la Investigación Clínica. | Temixco |
| Mercedes Paredes | JM Research SC | Cuernavaca |
| Pedro Sánchez | Instituto Nacional de Pediatría | Coyoacán |
| **Nepal** | | |
| Piush Kanodia | Nepalgunj Medical college Teaching Hospital | Nepalgunj |
| Dipesh Tamrakar | Dhulikhel Hospital, Kathmandu University Hospital | Dhulikhel |
| Santa Kumar Das | Institute of Medicine, Tribhuvan University | Kathmandu |
| **Uganda** | | |
| Patricia Nahirya Ntege | Baylor College of Medicine Children’s Foundation-Uganda | Kampala |
| Brenda Okech | Uganda Virus Research Institute (UVRI)-IAVI HIV Vaccine Program LTD. CRS | Entebbe |
| Hannah Kibuuka | Makerere University Walter Reed Project | Kampala |
| Cissy Kityo Mutuluuza | Joint Clinical Research Centre | Kampala |
| Anne Wajja | Medical Research Council/Uganda Virus Research Institute/London School of Hygiene and Tropical Medicine Uganda Research Unit | Entebbe |
| Deo Wabwire | MUJHU Research Collaboration / MUJHU Care Ltd | Kampala |
| Noah Kiwanuka | Africa Medical And Behavioural Sciences Organisation, Nansana | Waikso |
| Francis Kiweewa | Strengthening Institutional Capacity for Research Administration at Lira Regional Hospital | Lira |
| **Ukraine** | | |
| Svitlana Postol | Synexus Affiliate - MC of LLC Medbud-Clinic | Kyiv |
| Hanna Beyko | Center of Family Medicine Plus LLC | Kyiv |
| Oleksandra Bilotkach | AES - AS - Medical Center of Edelweiss Medics LLC | Kyiv |
| Dmytro Dobryanskyi | Medical Center Medical Clinic Blagomed LLC | Kyiv |

#### 1.2 Randomization and blinding

Randomization was conducted using an interactive response technology system and was stratified by age (18–59 years/≥60 years), baseline SARS-CoV-2 rapid serodiagnostic test positivity, and study site. All participants, outcome assessors and laboratory staff performing assays were blinded to group assignments; only site staff involved in the preparation and administration of the vaccines were unblinded, but they were not involved in study outcome assessments.

#### 1.3 Participant inclusion/exclusion criteria

Patients are eligible for the study only if all the following criteria are met:

- Aged ≥18 years on the day of inclusion
- No intention to receive an authorized/approved COVID-19 vaccine outside of the study
- SARS-CoV-2 rapid serodiagnostic test performed at the time of enrolment to detect presence of SARS-CoV-2 antibodies
- For persons living with HIV, their HIV infection must be stable as determined by ongoing anti-retroviral treatment and a CD4 cell count of >200 cells/mm^3^
- Females of childbearing potential must agree to use an effective contraceptive method or remain abstinent from ≥4 weeks prior to the first study intervention administration until ≥12 weeks after the second study intervention administration; have a negative highly sensitive pregnancy test ≤25 hours before any dose of study intervention
- Females of non-childbearing potential must be post-menopausal for ≥1 year or surgically sterile
- Provision of a signed and dated informed consent form
- Able to attend all visits and to comply with all study procedures
- Covered by health insurance, if required by local, regional or national regulations

Participants are not eligible for the study if any of the following criteria are met:

- Known systemic hypersensitivity to any of the vaccine components or a history of a life-threatening reaction to a vaccine containing any of the same substances
- Dementia or any other cognitive condition at a stage that could interfere with following the study procedures based on Investigator’s judgement
- Self-reported thrombocytopenia, contraindicating intramuscular vaccination based on Investigator’s judgement
- Bleeding disorder or receipt of anticoagulants in the past 21 days preceding inclusion, contraindicating intramuscular vaccination based on Investigator’s judgement
- Unstable acute or chronic illness that in the opinion of the Investigator or designee poses additional risk as a result of participation or that could interfere with the study procedures
- Moderate or severe acute illness/infection (according to investigator judgement) on the day of vaccination or febrile illness (temperature ≥38°C [≥100.4°F]). A prospective participant should not be included in the study until the condition has resolved or the febrile event has subsided
- Receipt of any vaccine in the 30 days preceding or on the day of the first study vaccination or planned receipt of any vaccine between the first study vaccination and in the 30 days following the second study vaccination except for influenza vaccination, which may be received at any time in relation to study intervention
- Prior administration of a coronavirus vaccine (SARS-CoV-2, SARS-CoV, MERS-CoV)
- Receipt of solid-organ or bone marrow transplants in the past 180 days
- Receipt of anti-cancer chemotherapy in the last 90 days
- Participation at the time of study enrolment (or in the 30 days preceding the first study vaccination) or planned participation during the present study period in another clinical study investigating a vaccine, drug, medical device or medical procedure
- Deprived of freedom by an administrative or court order, or in an emergency setting, or hospitalized involuntarily
- Identified as an Investigator or employee of the Investigator or study center with direct involvement in the proposed study, or identified as an immediate family member (i.e. parent, spouse, natural or adopted child) of the Investigator or employee with direct involvement in the proposed study

#### 1.4 Participant counselling on approved/authorized vaccines

Since approved/authorized COVID-19 vaccines were already available in some countries and regions where the study was conducted, investigators discussed their availability with participants, encouraged them to obtain the approved/authorized vaccine as applicable, and proceeded with enrollment only if, despite encouragement, the participant expressed no interest in seeking approved/authorized COVID-19 vaccines. Furthermore, participants were counselled at each opportunity about the availability and benefits of approved vaccines. Participants were allowed to receive an authorized vaccine outside the study protocol and were offered the option to continue in the study for safety and immunogenicity follow-ups.

#### 1.5 Participants at higher risk of severe COVID-19

High-risk conditions are conditions considered to be associated with an increased risk of severe COVID-19 (https://www.cdc.gov/coronavirus/2019-ncov/need-extra-precautions/evidence-table.html) and include:

- cancer
- chronic kidney disease
- chronic obstructive pulmonary disease (COPD)
- immunocompromised state from solid organ transplant
- immunocompromised state from other causes (blood or bone marrow transplant, immune deficiencies, HIV, use of corticosteroids, or use of immunosuppressors)
- obesity (body mass index of ≥30)
- heart conditions such as heart failure
- coronary artery disease or cardiomyopathies
- sickle cell disease
- thalassemia
- type 1 or type 2 diabetes mellitus
- moderate-to-severe asthma
- cerebrovascular disease
- cystic fibrosis
- hypertension/high blood pressure
- neurologic conditions
- hepatic disease
- pulmonary fibrosis
- smoking

#### 1.6 Injection protocol for the bivalent vaccine

The CoV2 preS dTM antigen and AS03 adjuvant were presented in two separate vials: a multi-dose vial containing AS03 (sufficient for 10 injections) multidose vial (3 mL target fill volume [20 mg/mL of D614 + 20 mg/mL B.1.351]). An equal volume of the adjuvant emulsion was added to the vial containing the antigen and mixed prior to injection.

Participants in the vaccine group received two 0.5 mL injections of the bivalent vaccine and participants in the placebo group received two 0.5 mL injections of 0.9% normal saline.

#### 1.7 Testing procedures and criteria for determination of prior SARS-CoV-2 infection

Participants were classified as naïve or non-naive at Day 1 and Day 22 or Day 1 or Day 22 by assessment of blood samples using Elecsys electrochemiluminescence immunoassays for detection of anti-S antibodies (Elecsys Anti-SARS-CoV-2 S assay; Roche, Indianapolis, IN, USA) on study Day 1 and for detection of anti-nucleocapsid antibodies (Elecsys Anti- SARS-CoV-2 N; Roche) on study Days 1 and 22; and detection of SARS-CoV-2 nucleic acids in nasopharyngeal swabs using nucleic-acid amplification tests (NAAT; Abbott RealTime SARS-CoV-2 assay; Abbott Molecular, Des Plaines, IL, USA) on study Days 1 and 22. Analyses were done according to the manufacturers’ instructions. Participants were defined as naive to SARS-CoV-2 on study Days 1 and 22 if they tested negative for anti-S antibodies on study Day 1 and for both anti-nucleocapsid antibodies and SARS-CoV-2 nucleic acids on Days 1 and 22; we defined participants as non-naive if they tested positive on at least one of the three tests on study Days 1, 22, or both.

The prior SARS-CoV-2 infection status of all randomized participants were defined in accordance with the below criteria.

| **Prior SARs-CoV-2 infection status** | **Description** |
| --- | --- |
| SARS-CoV-2 Naïve at baseline (Naïve-D1) | - Negative by the anti-S immunoassay (Roche Elecsys) on D01 serum sample   AND   - Negative by the anti-N immunoassay on D01 serum sample   AND   - Negative NAAT for SARS-CoV-2 on respiratory sample collected on D01 |
| SARS-CoV-2 Non-Naïve at baseline (Non-Naïve-D1) | - Positive by the anti-S immunoassay (Roche Elecsys) on D01 serum sample   OR   - Positive by the anti-N immunoassay on D01 serum sample   OR   - Positive NAAT for SARS-CoV-2 on respiratory sample collected on D01 |
| SARS-CoV-2 Naïve at second injection (Naïve-D1+D22) | - Negative by the anti-S immunoassay (Roche Elecsys) on D01 serum sample   AND   - Negative by anti-N immunoassay on D01 and D22 serum samples   AND   - Negative NAAT for SARS-CoV-2 on respiratory samples collected on D01 and D22 |
| SARS-CoV-2 Non-Naïve at second injection (Non-Naïve -D1/D22) | - Positive by the anti-S immunoassay (Roche Elecsys) on D01 serum sample   OR   - Positive by the anti-N immunoassay on D01 or D22 serum samples   OR   - Positive NAAT for SARS-CoV-2 on respiratory samples collected on D01 or D22 |

#### 1.8 Definition of COVID-19 like illness

New onset or exacerbation of any ONE of the following:

- Fever (measured temperature ≥ 100.4ºF OR ≥ 38.0ºC)
- Difficulty breathing or shortness of breath
- Altered level of consciousness
- Myocarditis, myocardial infarction
- Thromboembolic event (blood clots [eg, pulmonary embolism, deep vein thrombosis, stroke])
- Purpura fulminans
- Clinical or radiographic evidence of pneumonia
- Chilblains (COVID-toes)

OR

New onset or exacerbation of ANY ONE of the following (that persists for a period of at least 24 hours or reoccurs after a 12-hour period):

- Cough (dry or productive)
- Anosmia or partial loss of smell
- Ageusia or dysgeusia (loss or disturbance of taste)

OR

New onset of any TWO of the following symptoms that are present at the same time (both symptoms that persist for a period of at least 24 hours or reoccur after a 12-hour period):

- Sore throat
- Chills
- Myalgia
- Fatigue
- Malaise
- Headache
- Rhinorrhea or nasal congestion
- Abdominal pain
- At least one of nausea, diarrhea, vomiting

#### 1.9 Procedure in patients with laboratory confirmed NAAT and COVID-19 infection

Further anterior nasal swabs were collected 7–9 days and 12–14 days after the first illness visit. If any specimen was found to be positive for SARS-CoV-2, the participant was asked to continue recording their daily COVID-19 symptoms until the end of their illness or for up to 30 days from symptom onset. If symptoms persisted for over 30 days, participants were asked to record the date the symptoms resolved.

#### 1.10 Additional reported analyses and endpoints

The impact of sex, age (18–59 years and ≥60 years), and high-risk medical conditions on the above outcomes was evaluated as pre-defined exploratory objectives. Other analyses included the occurrence of symptomatic or severe COVID-19 ≥14 days after the first injection. The occurrence of asymptomatic infection in SARS-CoV2 naïve participants was also reported.

#### 1.11 Definition of COVID-19 efficacy outcomes

**Virologically-confirmed SARS-CoV-2 infection**

Defined as a positive result for SARS CoV-2 by NAAT on at least one respiratory sample. This includes positive results by any NAAT including tests performed outside the study protocol if confirmed by the adjudication committee.

**Symptomatic COVID-19**

Defined as virologically-confirmed SARS-CoV-2 infection accompanied by protocol-defined COVID-19-like illness.

**Asymptomatic SARS-CoV-2 infection**

Defined as SARS-CoV-2 infection, with no reported COVID-19-like illness episodes between enrolment and 14 days after the timepoint at which SARS-CoV-2 infection is ascertained, based on samples taken during previous visits.

**Hospitalized COVID-19**

Defined as an episode of Symptomatic COVID-19 that requires inpatient hospitalization.

**Moderate COVID-19**

Defined as Symptomatic COVID-19 with:

- Shortness of breath that persists for at least 12 hours

OR

- Clinical signs of moderate illness measured at least on two occasions separated by 30 mins (respiratory rate [RR] ≥ 20 breaths per minute at rest AND heart rate [HR] ≥ 90 beats per minute at rest)

AND

- No clinical signs indicative of severe COVID-19

**Severe COVID-19: defined as COVID-19 with any one of the following:**

- Any clinical signs of severe illness measured at least on 2 occasions separated by 30 minutes (saturation of oxygen [SpO2] ≤ 93% on room air (corrected for altitude), PaO2/FiO2 ˂ 300 mm Hg, RR ≥ 30 breaths per minute at rest, HR ≥ 125 beats per minute at rest)
- Supplemental oxygen administration for > 1 hour
- Use of invasive or non-invasive ventilation or Extracorporeal Membrane Oxygenation
- Clinical diagnosis of respiratory failure (ie, clinical need for one of the preceding therapies, but preceding therapies not able to be administered in setting of resource limitation)
- Significant acute renal, hepatic, or neurologic dysfunction
- Shock (defined by systolic blood pressure ˂ 90 mm Hg, or diastolic blood pressure ˂ 60 mm Hg or requiring vasopressors)
- Admission to an ICU
- Death

**COVID-19 severity scale**

The COVID-19 severity scale is based on the ordinal scale of clinical assessment:

1. Death

2. Hospitalized, on invasive mechanical ventilation or extracorporeal membrane oxygenation

3. Hospitalized, on non-invasive ventilation or high flow oxygen devices

4. Hospitalized, requiring supplemental oxygen

5. Hospitalized, not requiring supplemental oxygen – discharged but requiring ongoing medical care (COVID-19 related or otherwise)

6. Hospitalized, not requiring supplemental oxygen – discharged without ongoing medical care

7. Not hospitalized

**Death associated with COVID-19**

Death associated with COVID-19 is defined as death in a participant with COVID-19 who died within 28 days of the first positive specimen date if association confirmed by adjudication committee OR died more than 28 days after the first specimen date and COVID-19 is mentioned as an immediate or underlying cause of death on the death certificate if association confirmed by the adjudication committee.

#### 1.12 Safety data

Safety data included all immediate unsolicited systemic AEs within 30 minutes of each injection, medically attended AEs (MAAEs), serious AEs (SAEs), AEs of special interest (AESIs) including potential immune-mediated diseases (pIMDs).

AESIs included:

- Anaphylactic reactions
- Generalized convulsion
- Thrombocytopenia
- Thrombosis with Thrombocytopenia Syndrome*
- Myocarditis*
- Pericarditis*
- New onset and worsening of potential immune-mediated diseases^1^

*Due to safety signals detected after the use of other COVID-19 vaccine platforms from other manufacturers, Thrombosis with Thrombocytopenia Syndrome, Myocarditis and Pericarditis were also considered AESIs.

#### 1.13 Sample size calculations

It was estimated that a sample of 10,886 participants would be required, with approximately 125 symptomatic COVID-19 cases needed to achieve 80% statistical power. Participants were enrolled and randomized with allocation ratio (1:1) into the vaccine group and the placebo group. Among those, the planned target for SARS-CoV-2 non-naïve participants was approximately 3,266 participants (1,633 per arm). The sample size of 7,620 SARS-CoV-2 naïve participants was powered independently to demonstrate the primary objective of vaccine efficacy (VE) against symptomatic COVID-19 in SARS-CoV-2 naïve adults; however, only 1,176 were naïve at D1. Of note, the primary endpoint, including both naïve and non-naïve participants, was changed after enrollment was already completed; therefore, sample size calculations were based on a primary endpoint that considered only naïve participants. The power of primary efficacy analysis is driven by the total number of symptomatic COVID-19 events.

The incidence rate of symptomatic COVID-19 in Placebo is assumed as 2.25% illness rate per 2-months follow-up period. An attrition rate of 30% was expected, as, during the conduct of the study, a greater proportion of the cohort became eligible to receive locally available authorized COVID-19 vaccines.

The power of primary efficacy analysis was driven by the total number of symptomatic COVID-19 events. Because Omicron was the prevalent variant during case accrual for Stage 2 and the expected VE against Omicron was expected to be lower than the original assumption of 70%, the expected true VE for symptomatic COVID-19 for Stage 2 was estimated at 60%. Therefore, a total of approximately 125 symptomatic COVID-19 events was required to achieve 80% power with 1-side type I error rate of 0.025, assuming no interim analysis. For interim analyses, type I error rate was adjusted appropriately.

For the primary endpoint, the confidence intervals (CI) corresponding to vaccine efficacy estimates were calculated by an exact method,^2^ assuming a binomial distribution of the number of cases in the vaccine group, conditional to the total number of cases.

For other analyses, an unadjusted 95% CI was calculated.

#### 1.14 Descriptions of the analysis sets

| Population | Description |
| --- | --- |
| Screened | All participants screened for potential study enrolment will be included regardless of being enrolled or not being enrolled. The screening includes the SARS-CoV-2 rapid serodiagnosis test results, demographic information (age, ethnic/racial population, high-risk medical conditions), and inclusion/exclusion criteria. The participants reaching the enrolment cap identified in IRT will be excluded from the study enrolment and will have no participant ID assigned. |
| Randomized | All participants with a randomized group that has been allocated by IRT |
| Full analysis set (FAS) | All randomized participants who receive at least one study injection.  Participants will be analyzed according to the intervention to which they were randomized |
| Safety Analysis Set (SafAS) | All randomized participants who have received at least one dose of the study vaccine or placebo and with documented safety data.  All participants will have their safety analysed after each dose according to the intervention they actually received, and after any dose according to the intervention received at the first dose.  Safety data recorded for participants not administered a study intervention will be excluded from the analysis (and listed separately). |
| Reactogenicity Safety Analysis Subset (RsafAS) | Subset of the SafAS and comprising all participants who receive at least one study injection and are randomized into the reactogenicity subset and who have reported reactogenicity data. |
| Modified Full Analysis Set post-dose 1 (mFAS-PD1) | Subset of the FAS excluding:  • Participants with onset of symptomatic COVID-19 episode between the date of the first injection and 14 days after the first injection  • Participant discontinued from study before 14 days after the first injection |
| Modified Full Analysis Set post-dose 2 (mFAS-PD2) | Subset of the FAS excluding:   - Participants who did not complete the vaccination schedule (2 injections) - Participants with onset of symptomatic COVID-19 episode between the date of the first injection and before 14 days after the second injection - Participant received the second injection despite meeting any of the definitive contraindication criteria - Participant discontinued from study before 14 days after the second injection |

### 2.0 Supplementary efficacy data

#### 2.1 Reasons for discontinuation

|  | Vaccine group (N=6512) | Placebo group (N=6490) | All (N=13002) |
| --- | --- | --- | --- |
| **Overall (n=414)** | | | |
| Adverse event*, n (%) | 4 (<0.1) | 5 (<0.1) | 9 (<0.1) |
| Protocol deviation, n (%) | 26 (0.4) | 23 (0.4) | 49 (0.4) |
| Withdrawal by subject, n (%) | 146 (2.2) | 160 (2.2) | 306 (2.4) |
| Lost to follow-up, n (%) | 26 (0.4) | 24 (0.4) | 50 (0.4) |
| **Discontinued after second injection (n=89)** | | | |
| Adverse event*, n (%) | 1 (<0.1) | 3 (<0.1) | 4 (<0.1) |
| Protocol deviation, n (%) | 1 (<0.1) | 0 | 1 (<0.1) |
| Withdrawal by subject, n (%) | 29 (0.4) | 44 (0.7) | 73 (0.6) |
| Lost to follow-up, n (%) | 4 (<0.1) | 7 (0.1) | 11 (<0.1) |

*All adverse events were assessed as unrelated to the study intervention.

#### 2.2 Main analysis sets by age group, prior SARS-CoV-2 infection at baseline, and presence of high-risk medical conditions

|  | | **Vaccine group (N=6,512)**  **n** | **Placebo group (N=6,490)**  **n** |
| --- | --- | --- | --- |
| Overall | FAS | 6,472 | 6,452 |
|  | mFAS-PD2 | 5,736 | 5,680 |
|  | SafAS | 6,472 | 6,450 |
|  | RSafAS | 2,433 | 2,418 |
| 18–59 years | FAS | 6,078 | 6,069 |
|  | mFAS-PD2 | 5,404 | 5,350 |
|  | SafAS | 6,078 | 6,067 |
|  | RsafAS | 2,040 | 2,037 |
| ≥60 years | FAS | 394 | 383 |
|  | mFAS-PD2 | 332 | 330 |
|  | SafAS | 394 | 383 |
|  | RsafAS | 393 | 381 |
| Naïve at D1 | FAS | 588 | 588 |
|  | mFAS-PD1 | 577 | 576 |
|  | SafAS | 588 | 588 |
|  | RsafAS | 332 | 347 |
| Non-naïve at D1 | FAS | 4,860 | 4,833 |
|  | mFAS-PD1 | 4,826 | 4,793 |
|  | SafAS | 4,860 | 4,831 |
|  | RsafAS | 1,835 | 1,802 |
| Naïve at D1 and D22 | FAS | 333 | 350 |
|  | mFAS-PD2 | 327 | 343 |
| Non-naïve at D1 or D22 | FAS | 5,478 | 5,488 |
|  | mFAS-PD2 | 4,841 | 4,818 |
| Presence of high risk medical conditions | FAS | 2,095 | 2,070 |
|  | mFAS-PD2 | 1,793 | 1,786 |
|  | SafAS | 2,095 | 2,070 |
|  | RsafAS | 869 | 862 |
| No presence of high risk medical conditions | FAS | 4,377 | 4,382 |
|  | mFAS-PD2 | 3,943 | 3,894 |
|  | SafAS | 4,377 | 4,380 |
|  | RsafAS | 1,564 | 1,556 |

N=number of randomised participants.

Abbreviations: FAS, Full analysis set; mFAS-PD1, modified full analysis set post-dose 1; mFAS-PD2, modified full analysis set post-dose 2; RsafAS, reactogenicity safety analysis set; SafAS, safety analysis set.

The product received by two participants (i.e. vaccine or placebo) could not be determined; therefore, there is a difference of two participants between the FAS and the SafAS; the same difference is observed between the FAS and SafAS post-dose 1.

#### 2.3 High risk medical conditions - SafAS

|  | **Vaccine group (N=6,472) n (%)** | **Placebo group (N=6,450) n (%)** | **Total  (N=12,924) n (%)** |
| --- | --- | --- | --- |
| No | 4377 (67.6) | 4380 (67.9) | 8759 (67.8) |
| At least one high-risk medical condition | 2095 (32.4) | 2070 (32.1) | 4165 (32.2) |
| Smoking | 1068 (16.5) | 1040 (16.1) | 2108 (16.3) |
| Obesity (body mass index of 30 or higher) | 616 (9.5) | 580 (9.0) | 1196 (9.3) |
| Immunocompromised State from Other Causes | 323 (5.0) | 342 (5.3) | 665 (5.1) |
| Hypertension / High Blood Pressure | 281 (4.3) | 309 (4.8) | 590 (4.6) |
| Type 2 Diabetes Mellitus | 92 (1.4) | 84 (1.3) | 176 (1.4) |
| Moderate to Severe Asthma | 24 (0.4) | 22 (0.3) | 46 (0.4) |
| Chronic Kidney Disease | 11 (0.2) | 9 (0.1) | 20 (0.2) |
| Hepatic Disease | 11 (0.2) | 7 (0.1) | 18 (0.1) |
| Neurologic Conditions | 10 (0.2) | 8 (0.1) | 18 (0.1) |
| Type 1 Diabetes Mellitus | 8 (0.1) | 9 (0.1) | 17 (0.1) |
| Cardiovascular Disorder / Heart Failure | 7 (0.1) | 9 (0.1) | 16 (0.1) |
| Cancer | 7 (0.1) | 4 (<0.1) | 11 (<0.1) |
| Coronary Artery Disease or Cardiomyopathies | 8 (0.1) | 3 (<0.1) | 11 (<0.1) |
| Chronic Obstructive Pulmonary Disease | 6 (<0.1) | 4 (<0.1) | 10 (<0.1) |
| Immunocompromised State from Solid Organ Transplantation | 4 (<0.1) | 1 (<0.1) | 5 (<0.1) |
| Sickle Cell Disease | 3 (<0.1) | 0 | 3 (<0.1) |

#### 2.4 Follow-up data – SafAS (PD1)

|  | **Vaccine group** | **Placebo group** | **Total** |
| --- | --- | --- | --- |
| All |  |  |  |
| Follow-up duration,* days | 148 | 148 | 148 |
| Median subject  follow-up duration,^†^ days | 85 | 85 | 85 |
| Total follow-up duration,^‡^  1000-person years | 1.335 | 1.335 | – |
| Aged 18–25 years |  |  |  |
| Follow-up duration,* days | 147 | 148 | 148 |
| Median subject  follow-up duration,^†^ days | 79 | 83 | 82 |
| Total follow-up duration,^‡^  1000-person years | 0.320 | 0.322 | – |
| Aged 18–59 years |  |  |  |
| Follow-up duration,* days | 148 | 148 | 148 |
| Median subject  follow-up duration,^†^ days | 85 | 85 | 85 |
| Total follow-up duration,^‡^  1000-person years | 1.256 | 1.259 | – |
| Aged ≥60 years |  |  |  |
| Follow-up duration,* days | 124 | 125 | 125 |
| Median subject  follow-up duration,^†^ days | 76 | 71 | 72 |
| Total follow-up duration,^‡^  1000-person years | 0.079 | 0.076 | – |
| Aged 18–64 years |  |  |  |
| Follow-up duration,* days | 148 | 148 | 148 |
| Median subject  follow-up duration,^†^ days | 85 | 85 | 85 |
| Total follow-up duration,^‡^  1000-person years | 1.300 | 1.296 | – |
| Aged ≥65 years |  |  |  |
| Follow-up duration,* days | 124 | 124 | 124 |
| Median subject  follow-up duration,^†^ days | 76 | 76 | 76 |
| Total follow-up duration,^‡^  1000-person years | 0.034 | 0.039 | – |

*Follow-up duration: data cut-off date – minimum of all SafAS subjects (Visit 01 date) + 1.
^†^Subject follow-up duration: minimum (data cut-off date, termination date or death date) – Visit 01 date + 1.
^‡^Total study follow-up duration: sum of subject follow-up duration / (365.25 * 1000)

#### 2.5 Follow-up data – mFAS-PD2

|  | **Vaccine group** | **Placebo group** | **Total** |
| --- | --- | --- | --- |
| All |  |  |  |
| Follow-up duration,* days | 118 | 118 | 118 |
| Median subject  follow-up duration,^†^ days | 58 | 58 | 58 |
| Total follow-up duration,^‡^  1000-person years | 0.824 | 0.818 | – |
| Aged 18–25 years |  |  |  |
| Follow-up duration,* days | 117 | 118 | 118 |
| Median subject  follow-up duration,^†^ days | 56 | 57 | 56 |
| Total follow-up duration,^‡^  1000-person years | 0.191 | 0.192 | – |
| Aged 18–59 years |  |  |  |
| Follow-up duration,* days | 118 | 118 | 118 |
| Median subject  follow-up duration,^†^ days | 58 | 58 | 58 |
| Total follow-up duration,^‡^  1000-person years | 0.779 | 0.773 | – |
| Aged ≥60 years |  |  |  |
| Follow-up duration,* days | 100 | 104 | 104 |
| Median subject  follow-up duration,^†^ days | 54 | 50 | 51 |
| Total follow-up duration,^‡^  1000-person years | 0.045 | 0.045 | – |
| Aged 18–64 years |  |  |  |
| Follow-up duration,* days | 118 | 118 | 118 |
| Median subject  follow-up duration,^†^ days | 58 | 58 | 58 |
| Total follow-up duration,^‡^  1000-person years | 0.803 | 0.795 | – |
| Aged ≥65 years |  |  |  |
| Follow-up duration,* days | 100 | 100 | 100 |
| Median subject  follow-up duration,^†^ days | 56 | 51 | 55 |
| Total follow-up duration,^‡^  1000-person years | 0.021 | 0.023 | – |

*Follow-up duration: Data cut-off date - Minimum of all mFAS-PD2 subjects (date of V02) + 1. ^†^Subject follow-up duration: Minimum (data cut-off date, termination date, or death date) - V02 date + 1. ^‡^Total follow-up duration: Sum of subject follow-up duration / (365.25 * 1000

#### 2.6 Vaccine efficacy - Sensitivity analysis including Ukrainian participants

|  | Vaccine group (N=5876) | | | Placebo group (N=5802) | | | Vaccine efficacy | |
| --- | --- | --- | --- | --- | --- | --- | --- | --- |
|  | Cases | 1000 person years at risk | Incidence rate (95% CI) | Cases | 1000 person years at risk | Incidence rate (95% CI) | % | 95% CI |
| Symptomatic COVID-19 | 33 | 0.609 | 54.22 (37.32; 76.14) | 90 | 0.597 | 150.75 (95% CI 121.25; 185.30) | 64.0 | 45.9; 76.6) |

Cases: number of subjects with ≥1 occurrence of relevant endpoint from 14 days post-injection 1 in the analysis population

#### 2.7 Vaccine efficacy in the mFAS-PD1 subset

|  | Vaccine group (n=6,418) | Placebo group (n=6,391) | Vaccine efficacy,  % (95% CI) |
| --- | --- | --- | --- |
| Symptomatic COVID-19 |  |  |  |
| Cases | 68 | 169 | – |
| 1000 person-years at risk | 1.069 | 1.055 | – |
| Incidence rate  (95% CI) | 63.593  (49.38–80.62) | 160.144  (136.91–186.19) | 60.3  (47.1–70.5) |
| Participants at risk, n | 6,418 | 6,390 | – |
| Cumulative incidence,  % (95% CI) | 1.1  (0.8–1.3) | 2.6  (2.3–3.1) | 59.9  (46.6–70.2) |
| Severe COVID-19 |  |  |  |
| Cases | 6 | 8 | – |
| 1000 person-years at risk | 1.069 | 1.055 | – |
| Incidence rate  (95% CI) | 5.611  (2.06–12.21) | 7.581  (3.27–14.94) | 26.0  (-143.3–78.8) |
| Participants at risk, n | 6,418 | 6,390 | – |
| Cumulative incidence,  % (95% CI) | 0.1 (0–0.2) | 0.1 (0.1–0.2) | 25.3  (-145.4–78.6) |

All cases correspond to one episode.
Incidence rate: cases per 1000 person-years at risk
Subjects at risk: subjects with censor date after 14 days post-injection 1 in the analysis population
Cumulative incidence: cases divided by number of subjects at risk in each group * 100

#### 2.8 Kaplan-Meier cumulative incidence of symptomatic COVID-19 in the mFAS-PD1 population (overall, naïve and non-naïve populations)

A. Overall

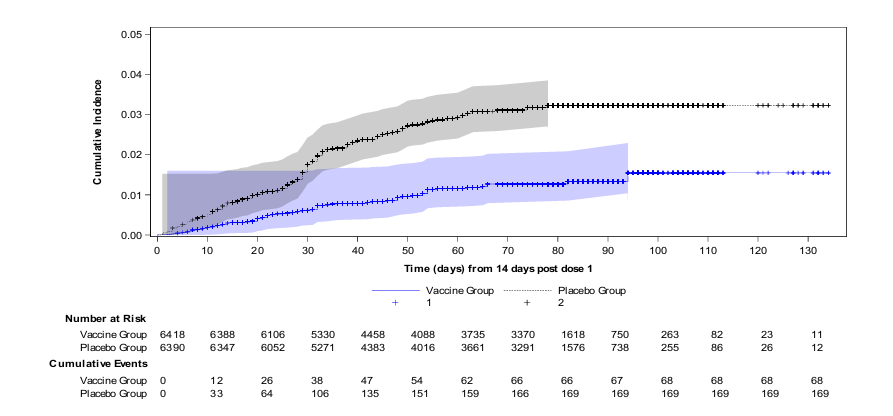

B. Naïve at baseline

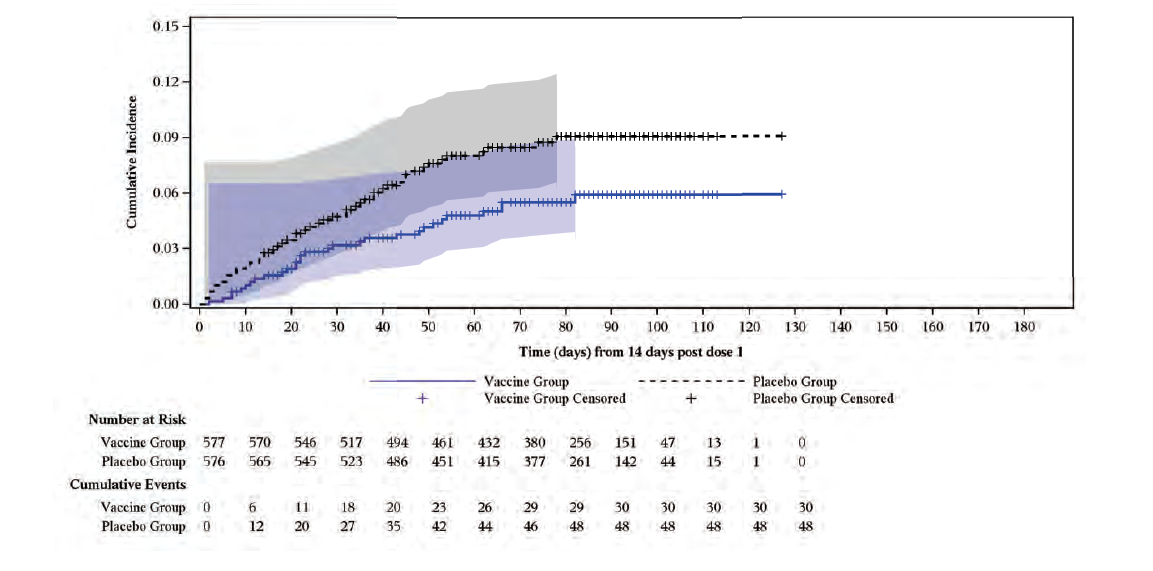

C. Non-naïve at baseline

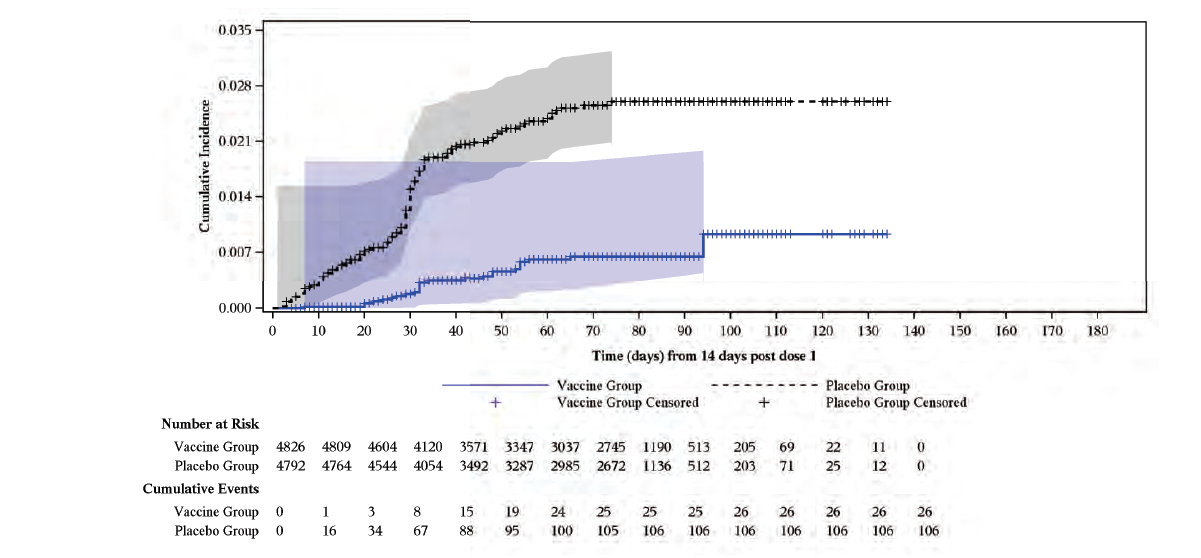

#### 2.9 Vaccine efficacy in the mFAS-PD2 population according to country, race/ethnic subgroups and risk of severe disease

|  | | Vaccine group (n=5,736) | Placebo group (n=5,680) | Vaccine efficacy,  % (95% CI) |
| --- | --- | --- | --- | --- |
| **Country** | **Colombia** |  |  |  |
|  | Cases | 3 | 7 | – |
|  | 1000 person-years at risk | 0.020 | 0.018 | – |
|  | Incidence rate  (95% CI) | 149.938 (30.92–438.18) | 385.459 (154.97–795.19) | 61.1  (-70.4–93.5) |
|  | Participants at risk, n | 328 | 319 | – |
|  | Cumulative incidence,  % (95% CI) | 0.9 (0.2–2.6) | 2.2 (0.9–4.5) | 58.3  (-82.6–93.0) |
|  | **Ghana** |  |  |  |
|  | Cases | 0 | 1 | – |
|  | 1000 person-years at risk | 0.007 | 0.068 | – |
|  | Incidence rate  (95% CI) | 0 (0–558.38) | 14.635  (0.37–81.54) | 100 (-3614.1–100.0) |
|  | Participants at risk, n | 570 | 548 | – |
|  | Cumulative incidence,  % (95% CI) | 0 (0–0.6) | 0.2 (0–1.0) | 100 (-3649.5–100.0) |
|  | **India** |  |  |  |
|  | Cases | 6 | 29 | – |
|  | 1000 person-years at risk | 0.199 | 0.196 | – |
|  | Incidence rate  (95% CI) | 30.188  (11.08–65.71) | 148.129 (99.2–212.74) | 79.6  (50.0–93.1) |
|  | Participants at risk, n | 1628 | 1618 | – |
|  | Cumulative incidence,  % (95% CI) | 0.4 (0.1–0.8) | 1.8 (1.2–2.6) | 79.4  (49.6–93.0) |
|  | **Kenya** |  |  |  |
|  | Cases | 3 | 7 | – |
|  | 1000 person-years at risk | 0.151 | 0.149 | – |
|  | Incidence rate  (95% CI) | 19.915  (4.11–58.20) | 47.042  (18.91–96.93) | 57.7  (-85.4–92.9) |
|  | Participants at risk, n | 1620 | 1614 |  |
|  | Cumulative incidence,  % (95% CI) | 0.2 (0–0.5) | 0.4 (0.2–0.9) | 57.3  (-87.0–92.9) |
|  | **Mexico** |  |  |  |
|  | Cases | 18 | 31 | – |
|  | 1000 person-years at risk | 0.042 | 0.040 | – |
|  | Incidence rate  (95% CI) | 432.590 (256.38–683.68) | 772.357 (524.78–1096.30) | 44.0  (-3.3–70.5) |
|  | Participants at risk, n | 320 | 317 | – |
|  | Cumulative incidence,  % (95% CI) | 5.6 (3.4–8.7) | 9.8 (6.7–13.6) | 42.5  (-6.1–69.7) |
|  | **Nepal** |  |  |  |
|  | Cases | 2 | 14 | – |
|  | 1000 person-years at risk | 0.115 | 0.115 | – |
|  | Incidence rate  (95% CI) | 17.400  (2.11–62.86) | 121.860 (66.62–204.46) | 85.7  (37.8–98.4) |
|  | Participants at risk, n | 738 | 760 | – |
|  | Cumulative incidence,  % (95% CI) | 0.3 (0–1.0) | 1.8 (1.0–3.1) | 85.3  (35.9–98.4) |
|  | **Uganda** |  |  |  |
|  | Cases | 0 | 0 | – |
|  | 1000 person-years at risk | 0.007 | 0.007 | – |
|  | Incidence rate  (95% CI) | 0 (0–558.38) | 0 (0–545.05) | Not computed |
|  | Participants at risk, n | 107 | 2376 | – |
|  | Cumulative incidence,  % (95% CI) | 0 (0–3.4) | 1.8 (1.3–2.4) | Not computed |
| **Race/ Ethnicity** | **Asian** |  |  |  |
|  | Cases | 8 | 42 | – |
|  | 1000 person-years at risk | 0.313 | 0.310 | – |
|  | Incidence rate  (95% CI) | 25.539 (11.03–50.32) | 135.278 (97.50–182.86) | 81.1  (59.3–92.3) |
|  | Participants at risk, n | 2363 | 2376 | – |
|  | Cumulative incidence,  % (95% CI) | 0.3 (0.1–0.7) | 1.8 (1.3–2.4) | 80.8  (58.7–92.2) |
|  | **Black African or  African American** |  |  |  |
|  | Cases | 3 | 8 | – |
|  | 1000 person-years at risk | 0.228 | 0.223 | – |
|  | Incidence rate  (95% CI) | 13.134 (2.71–38.38) | 35.862 (15.48–70.66) | 63.4  (-52.6–93.7) |
|  | Participants at risk, n | 2295 | 2268 | – |
|  | Cumulative incidence,  % (95% CI) | 0.1 (0–0.4) | 0.4 (0.2–0.7) | 62.9  (-54.4–93.7) |
| **At risk of severe COVID-19*** | **Aged 18–59 years** |  |  |  |
|  | Cases | 10 | 28 | – |
|  | 1000 person-years at risk | 0.164 | 0.156 | – |
|  | Incidence rate  (95% CI) | 60.829 (29.17–111.87) | 179.727 (119.43–259.76) | 66.2  (28.2–85.3) |
|  | Participants at risk, n | 1429 | 1431 | – |
|  | Cumulative incidence,  % (95% CI) | 0.7 (0.3–1.3) | 2.0 (1.3–2.8) | 64.2 (24.1–84.5) |
|  | **Aged ≥65 years** |  |  |  |
|  | Cases | 1 | 0 | – |
|  | 1000 person-years at risk | 0.009 | 0.009 | – |
|  | Incidence rate  (95% CI) | 110.883 (2.81–617.80) | 0 (0–409.53) | Not computed |
|  | Participants at risk, n | 83 | 82 | – |
|  | Cumulative incidence,  % (95% CI) | 1.2 (0–6.5) | 0 (0–4.4) | Not computed |

*Comorbidities associated with increased risk of severe COVID-19 are listed above. All cases correspond to one episode. Incidence rate” is the “incidence expressed by 1000 patient-years at risk.

#### 2.10 Vaccine efficacy against asymptomatic SARS-CoV-2 infection in naïve participants

|  | mFAS-PD1 Naïve-D1 | | | mFAS-PD2 Naïve-D1+D22 | | |
| --- | --- | --- | --- | --- | --- | --- |
|  | Vaccine group (n=577) | Placebo group (n=576) | Vaccine efficacy,  %  (95% CI) | Vaccine group (n=315) | Placebo group (n=333) | Vaccine efficacy,  %  (95% CI) |
| Cases | 275 | 266 |  | 100 | 107 |  |
| Cumulative incidence,  % (95% CI) | 47.7 (43.5–51.8) | 46.2 (42.1–50.3) | –3.2  (-22.6–13.1) | 31.7 (26.6–37.2) | 32.1 (27.1–37.4) | 1.2  (-31.0–25.5) |

Cases: number of subjects with ≥1 occurrence of relevant endpoint from 14 days post-injection (1 or 2 as applicable) in the analysis population.

#### 2.11 Reasons for missing sequencing data in the adjudicated cases

Cases being diagnosed using a local test for which no specimen was available (n=8), the viral load threshold was too low for detection (n=12), the laboratory did not produce a valid result (n=32), or the sample was not tested (n=1). Of the 53 cases with undiagnosed variants, 18 out of 32 (56%) were in the vaccine group and 35 out of 89 (39%) were in the placebo group.

#### 2.12 Cumulative incidence of symptomatic Omicron infections in the mFAS-PD2 set

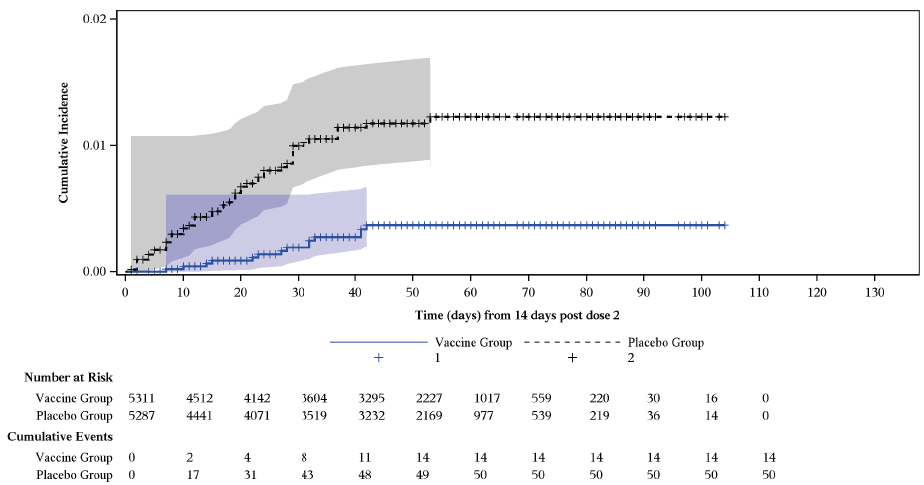

#### 2.13 Sensitivity analysis: efficacy against Symptomatic COVID-19 caused by Omicron or undefined variants in the mFAS-PD2 population

|  | Vaccine group  (n=5,736) | | | Placebo group  (n=5,680) | | | Vaccine efficacy, %  (95% CI) |
| --- | --- | --- | --- | --- | --- | --- | --- |
|  | Cases | n at risk | Cumulative incidence (95% CI) | Cases | n at risk | Cumulative incidence (95% CI) |  |
| All participants | 32 | 5,311 | 0.6  (0.4; 0.8) | 85 | 5,287 | 1.6 (1.3; 2.0) | 63.1  (43.9; 76.2) |
| Naïve participants | 15 | 315 | 4.8  (2.7; 7.7) | 21 | 333 | 6.3  (3.9; 9.5) | 27.6  (-47.3; 65.3) |
| Non-naïve participants | 16 | 4,519 | 0.4  (0.2; 0.6) | 60 | 4,514 | 1.3 (1.0; 1.7) | 73.8 (53.9; 85.9) |

Cases: number of participants with ≥1 symptomatic COVID-19 episode from 14 days post-injection 2 in the analysis population
Cumulative incidence: cases divided by the number of participants in each group * 100

#### 2.14 Safety overview after injection 1 and injection 2 in all participants – RsafAS/SafAS

|  | Post injection 1 | | | | | | Post injection 2 | | | | | |
| --- | --- | --- | --- | --- | --- | --- | --- | --- | --- | --- | --- | --- |
|  | Vaccine group (N=6472) | | | Placebo group (N=6450) | | | Vaccine group (N=5788) | | | Placebo group (N=5755) | | |
| Participants experiencing at least one: | n/M | % | 95% CI | n/M | % | 95% CI | n/M | % | 95% CI | n/M | % | 95% CI |
| SafAS |  |  |  |  |  |  |  |  |  |  |  |  |
| Within 30 minutes after injection |  |  |  |  |  |  |  |  |  |  |  |  |
| Immediate unsolicited AE | 4/6472 | <0.1 | (0; 0.2) | 4/6450 | <0.1 | (0; 0.2) | 0/5788 | 0 | (0; 0.1) | 3/5755 | <0.1 | (0; 0.2) |
| Immediate unsolicited AR | 4/6472 | <0.1 | (0; 0.2) | 3/6450 | <0.1 | (0; 0.1) | 0/5788 | 0 | (0; 0.1) | 3/5755 | <0.1 | (0; 0.2) |
| RSafAS |  |  |  |  |  |  |  |  |  |  |  |  |
| Solicited reaction within solicited  period after injection |  |  |  |  |  |  |  |  |  |  |  |  |
| Solicited reaction | 1119/2420 | 46.2 | (44.2; 48.3) | 685/2403 | 28.5 | (26.7; 30.4) | 911/2271 | 40.1 | (38.1; 42.2) | 629/2259 | 27.8 | (26.0; 29.7) |
| Grade 3 solicited reaction | 124/2420 | 5.1 | (4.3; 6.1) | 82/2403 | 3.4 | (2.7; 4.2) | 110/2271 | 4.8 | (4.0; 5.8) | 57/2259 | 2.5 | (1.9; 3.3) |
| Solicited injection site reaction | 862/2419 | 35.6 | (33.7; 37.6) | 412/2403 | 17.1 | (15.7; 18.7) | 720/2270 | 31.7 | (29.8; 33.7) | 376/2258 | 16.7 | (15.1; 18.3) |
| Grade 3 solicited injection site  reaction | 58/2419 | 2.4 | (1.8; 3.1) | 30/2403 | 1.2 | (0.8; 1.8) | 54/2270 | 2.4 | (1.8; 3.1) | 20/2258 | 0.9 | (0.5; 1.4) |
| Solicited systemic reaction | 818/2420 | 33.8 | (31.9; 35.7) | 560/2403 | 23.3 | (21.6; 25.0) | 690/2271 | 30.4 | (28.5; 32.3) | 520/2259 | 23.0 | (21.3; 24.8) |
| Grade 3 solicited systemic reaction | 104/2420 | 4.3 | (3.5; 5.2) | 75/2403 | 3.1 | (2.5; 3.9) | 91/2271 | 4.0 | (3.2; 4.9) | 52/2259 | 2.3 | (1.7; 3.0) |
| Within 21 days after injection |  |  |  |  |  |  |  |  |  |  |  |  |
| Unsolicited AE | 97/2433 | 4.0 | (3.2; 4.8) | 113/2418 | 4.7 | (3.9; 5.6) | 78/2276 | 3.4 | (2.7; 4.3) | 99/2271 | 4.4 | (3.6; 5.3) |
| Unsolicited AR | 14/2433 | 0.6 | (0.3; 1.0) | 11/2418 | 0.5 | (0.2; 0.8) | 13/2276 | 0.6 | (0.3; 1.0) | 5/2271 | 0.2 | (0.1; 0.5) |
| Unsolicited non-serious AE | 90/2433 | 3.7 | (3.0; 4.5) | 108/2418 | 4.5 | (3.7; 5.4) | 75/2276 | 3.3 | (2.6; 4.1) | 96/2271 | 4.2 | (3.4; 5.1) |
| Unsolicited non-serious AR | 14/2433 | 0.6 | (0.3; 1.0) | 11/2418 | 0.5 | (0.2; 0.8) | 13/2276 | 0.6 | (0.3; 1.0) | 5/2271 | 0.2 | (0.1; 0.5) |
| Unsolicited non-serious injection  site AR | 3/2433 | 0.1 | (0; 0.4) | 0/2418 | 0 | (0; 0.2) | 1/2276 | <0.1 | (0; 0.2) | 0/2271 | 0 | (0; 0.2) |
| Unsolicited non-serious systemic  AE | 87/2433 | 3.6 | (2.9; 4.4) | 108/2418 | 4.5 | (3.7; 5.4) | 74/2276 | 3.3 | (2.6; 4.1) | 96/2271 | 4.2 | (3.4; 5.1) |
| Participants experiencing at least one: |  |  |  |  |  |  |  |  |  |  |  |  |
| Unsolicited non-serious systemic  AR | 11/2433 | 0.5 | (0.2; 0.8) | 11/2418 | 0.5 | (0.2; 0.8) | 12/2276 | 0.5 | (0.3; 0.9) | 5/2271 | 0.2 | (0.1; 0.5) |
| SafAS |  |  |  |  |  |  |  |  |  |  |  |  |
| Within 21 days after injection |  |  |  |  |  |  |  |  |  |  |  |  |
| AE leading to study discontinuation | 2/6472 | <0.1 | (0; 0.1) | 0/6450 | 0 | (0; 0.1) | 0/5788 | 0 | (0; 0.1) | 2/5755 | <0.1 | (0; 0.1) |
| SAE | 11/6472 | 0.2 | (0.1; 0.3) | 8/6450 | 0.1 | (0.1; 0.2) | 6/5788 | 0.1 | (0; 0.2) | 6/5755 | 0.1 | (0; 0.2) |
| Death | 1/6472 | <0.1 | (0; 0.1) | 0/6450 | 0 | (0; 0.1) | 0/5788 | 0 | (0; 0.1) | 2/5755 | <0.1 | (0; 0.1) |
| AESI (including pIMDs) | 1/6472 | <0.1 | (0; 0.1) | 1/6450 | <0.1 | (0; 0.1) | 0/5788 | 0 | (0; 0.1) | 0/5755 | 0 | (0; 0.1) |
| MAAE | 169/6472 | 2.6 | (2.2; 3.0) | 172/6450 | 2.7 | (2.3; 3.1) | 101/5788 | 1.7 | (1.4; 2.1) | 112/5755 | 1.9 | (1.6; 2.3) |

Abbreviations: AE: adverse event; AESI: Adverse Event of Special Interest; AR: adverse reaction; MAAE: Medically-Attended Adverse Event; pIMDs: potential immune-mediated diseases; SAE: Serious Adverse Event

n: number of participants experiencing the endpoint; M: number of participants with available data for the relevant endpoint

Related SAEs: causal relationship reported by an investigator as related. If the relationship was missing, the SAE was considered as related

#### 2.15 Additional safety outcomes

- Observed SISRs and SSRs were mostly mild to moderate in severity, started within 3 days after the injection and spontaneously resolved within 1–3 days. The type, incidence and severity of SISRs and SSRs were comparable after both injections, irrespective of participant age.
- Ten (<0.1%) participants (five [<0.1%] in the Vaccine Group and five [<0.1%] in the Placebo Group), reported discontinuation from the study due to an AE; all AEs were assessed as not related to the study intervention.
- Up to the analysis cut-off date, 49 participants reported a pregnancy. Based on limited data (pregnancy precluded eligibility to the study), no safety concern was identified among pregnant women.
- There was no evidence of an increased risk of severe COVID-19 in the vaccine group compared with the placebo group. There were no fatal COVID-19 cases and a limited number of severe COVID-19 and hospitalized COVID-19 cases.

**References**

1.	Tavares Da Silva F, De Keyser F, Lambert P-H, Robinson WH, Westhovens R, Sindic C. Optimal approaches to data collection and analysis of potential immune mediated disorders in clinical trials of new vaccines. Vaccine 2013;31:1870-6.

2.	Breslow NE, Day NE. Statistical methods in cancer research. Volume II--The design and analysis of cohort studies. IARC scientific publications 1987:1-406.
